# Tool to assess risk of bias due to missing evidence in network meta-analysis (ROB-MEN): elaboration and examples

**DOI:** 10.1101/2021.05.02.21256160

**Authors:** Virginia Chiocchia, Adriani Nikolakopoulou, Julian PT Higgins, Matthew J Page, Theodoros Papakonstantinou, Andrea Cipriani, Toshi A Furukawa, George CM Siontis, Georgia Salanti

## Abstract

Selective outcome reporting and publication bias threaten the validity of systematic reviews and meta-analysis and ultimately can affect clinical decision-making. A rigorous methodology to evaluate the impact of this bias on the meta-analysis results of a network of interventions is still lacking. We present a tool to assess the Risk Of Bias due to Missing Evidence in Network meta-analysis (ROB-MEN) by expanding the methods previously developed for pairwise meta-analysis (ROB-ME, http://www.riskofbias.info).

ROB-MEN first evaluates the risk of bias due to missing evidence for each pairwise comparison separately. This step considers possible bias due to the presence of studies with unavailable results (*known unknowns*) and the potential for unpublished studies (*unknown unknowns*). The second step combines the overall judgements about the risk of bias due to missing evidence in pairwise comparisons with the percentage contribution of direct comparisons on the NMA estimates, the presence or absence of small-study effects, as evaluated by network meta-regression, and any bias from unobserved comparisons. Then, a level of “low risk”, “some concerns” or “high risk” for the bias due to missing evidence is assigned to each NMA estimate, which is our tool’s final output.

We describe the methodology of ROB-MEN step-by-step using an illustrative example from a published NMA of non-diagnostic modalities for the detection of coronary artery disease in patients with low risk acute coronary syndrome. We also report a full application of the tool on a larger and more complex published network of 18 drugs from head-to-head studies for the acute treatment of adults with major depressive disorder. The ROB-MEN tool is the first tool for evaluating the risk of bias due to missing evidence in NMA and it is applicable to networks of all sizes and geometry.

## 1 Introduction

One of the most challenging issues in evidence-based medicine is the bias introduced by the selective non-reporting of primary studies or results. Failure to report all findings can lead to results being missing from a meta-analysis; this can either be due to a whole study being missing, commonly referred to as ‘publication bias’, or because specific outcome results are not reported in a publication, usually referred to as ‘selective outcome reporting bias’ or ‘selective non-reporting of results’.

Several methods are available to investigate such bias in pairwise meta-analysis. These include generic approaches, for example, comparisons of study protocols with published reports and comparison of results obtained from published versus unpublished sources, as well as statistical methods (e.g. funnel plots [1–3], tests for small-study effects [1,4–6] and selection models [7,8]). Recently, a tool to evaluate Risk Of Bias due to Missing Evidence (ROB-ME) in pairwise meta-analysis has been presented [9]. ROB-ME involves several steps starting with the selection of the syntheses to be assessed for risk of bias due to missing evidence. The procedure then continues by identifying any studies with unavailable results (‘known unknowns’) and considering the potential for unpublished studies (‘unknown unknowns’) before reaching an overall judgement about the risk of bias due to missing evidence in each synthesized result (see Glossary of definitions, Box 2). The various approaches for assessing risk of bias due to missing results have been reviewed and described extensively [10,11].

Several of the approaches to evaluate or minimize bias developed for pairwise meta-analysis apply equally to network meta-analysis (NMA). For example, comparison of published and unpublished data for the same study is feasible and useful with any type of data synthesis. Several numerical approaches have been adapted to the NMA setting [12–16]. However, a rigorous methodology for assessing risk of bias due to missing results in NMA estimates is currently lacking.

To address this gap, we developed a tool for the assessment of bias due to missing evidence in NMA. We call this tool Risk Of Bias due to Missing Evidence in Network meta-analysis (ROB-MEN). We assume that investigators made their best efforts to assemble studies into a connected and coherent network according to a protocol, checked the assumptions of synthesis and deemed them plausible, and finally synthesized the study results using appropriate statistical methods to obtain all relative treatment effects between all pairs of interventions. Then, ROB-MEN can be used to assess the risk of bias due to missing evidence in each of the relative treatment effects as estimated in NMA.

In subsequent sections we explain the ROB-MEN approach step by step. In each step, we illustrate the new methodology using an example from a published NMA. Furthermore, after describing the methods we report a full application of the ROB-MEN tool in a network of 18 antidepressants from head-to-head studies [17].

### Illustrative example

#### Non-invasive diagnostic modalities for the detection of coronary artery disease in patients with low-risk acute coronary syndrome

To illustrate the steps, we use a network of six non-invasive diagnostic modalities for the detection of coronary artery disease in patients with low risk acute coronary syndrome (ACS) as previously reported by Siontis and others[18]. The outcome of interest is referral to invasive coronary angiography (ICA) and the diagnostic modalities are exercise electrocardiogram (ECG), single photon emission computed tomography-myocardial perfusion imaging (SPECT-MPI), coronary computed tomographic angiography (CCTA), cardiovascular magnetic resonance (CMR), stress echocardiography (stress echo) and standard care (based on the discretion of the clinicians and on locally applied diagnostic strategies). In Box 1 we show the network graph and summarize the analysis and results from NMA. These found that an initial diagnostic strategy of stress echo, CMR or exercise ECG is associated with fewer referrals for downstream invasive coronary angiography than non-invasive anatomical testing (CCTA). It also showed marginal differences, although more precise, for SPECT-MPI and standard care versus CCTA. We would like to make statements about the risk of bias due to missing evidence for each one of the 15 relative treatment effects.

##### Box 1

**Network graph, methods and forest plot for the network meta-analysis of non-invasive diagnostic modalities for the detection of coronary artery disease in patients with low risk acute coronary syndromes used as illustrative example**. ECG: electrocardiogram; SPECT-MPI: single photon emission computed tomography-myocardial perfusion imaging; CCTA: coronary computed tomographic angiography; CMR: cardiovascular magnetic resonance; Echo: echocardiography.

The network was reanalysed by fitting Bayesian random-effects models for network meta-analysis using the BUGSnet package in R. Summary odds ratios (ORs) and 95% credible intervals (CI) were estimated from binomial likelihoods models with common heterogeneity using an independent normal prior distribution with mean 0 and standard deviation 15*u* for the treatment effect and a uniform distribution with range 0 to *u* for the heterogeneity, where *u* represents the largest maximum likelihood estimator in single trials, as recommended by van Valkenhoef et al (*Res. Syn. Meth*. 2012, 3(4):285-99). The adjusted OR are estimated from a network meta-regression model using the smallest observed variance as a covariate and assuming unrelated coefficients. The prespecified prior for the unrelated regression coefficients is a *t*(0.*u*^2^.1) where *u* is again the largest maximum likelihood estimator in single trials. As reported in the original publication by Siontis et al. (*BMJ* 2018, 360:k504), there was no evidence of major inconsistency.

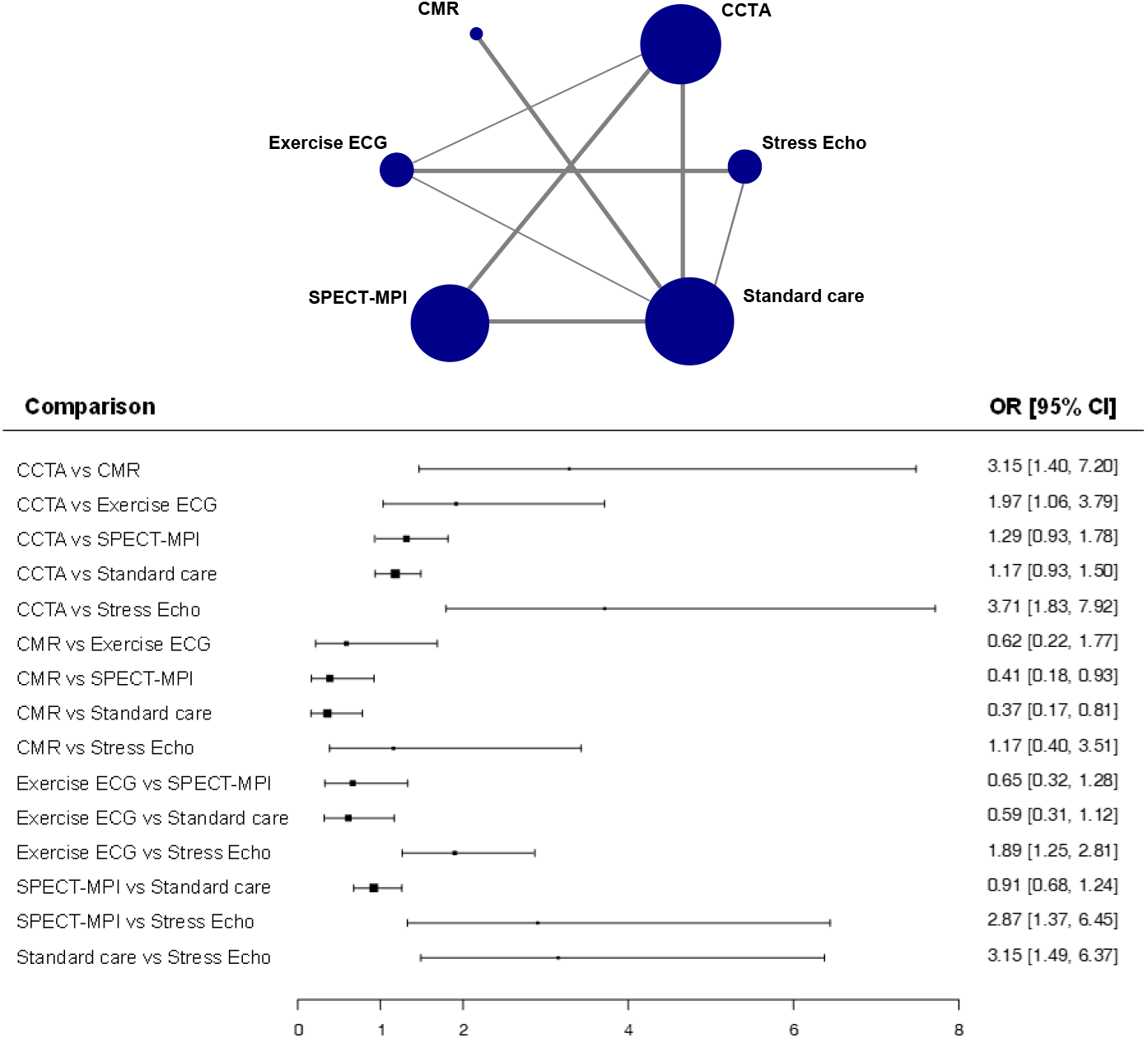

*Comparisons with direct evidence*: CCTA vs exercise ECG, CCTA vs SPECT-MPI, CCTA vs standard care, CMR vs standard care, exercise ECG vs standard care, exercise ECG vs stress echo, SPECT-MPI vs standard care, standard care vs stress echo.

*Comparisons with indirect evidence*: CCTA vs CMR, CCTA vs stress echo, CMR vs exercise ECG, CMR vs SPECT-MPI, CMR vs stress echo, exercise ECG vs SPECT-MPI, SPECT-MPI vs stress echo.

##### Box 2

**Glossary of terms as used in our manuscript**

**Pairwise comparisons:** all treatment comparisons in the network irrespective of the availability of data. A network with T treatments has T(T-1)/2 pairwise comparisons. Depending on whether there are studies reporting the studied outcome, the pairwise comparisons can be distinguished into *observed for this outcome, observed for other outcomes*, and *unobserved*.

**Direct evidence:** The evidence available (statistical information derived from data) about a pairwise comparison that is available from direct, within-study information about that comparison.

**Indirect evidence:** The evidence available (statistical information derived from data) about a pairwise comparison that is *not* available from within-study information, i.e. is obtained indirectly via a common comparator or chain of comparisons.

**‘Only direct’ estimate:** Relative treatment effect estimated in an NMA that is derived only from direct evidence.

**‘Only indirect’ estimate:** Relative treatment effect estimated in an NMA that is derived only from indirect evidence.

**Mixed estimate:** Relative treatment effect estimated in an NMA that is derived from both direct and indirect evidence.

**NMA estimate:** estimates of relative treatment effects derived from network meta-analysis; these can be distinguished into ‘Only direct’, ‘Only indirect’ and Mixed estimates.

**Known unknown bias:** bias arising from missing results due to selective outcome reporting i.e. results being reported, but not others, within studies published or otherwise known to exist.

**Unknown unknown bias:** bias introduced from missing studies because they are entirely unpublished i.e. not known to exist.

## 2 Methods

### 2.1 Overview of the ROB-MEN

In ROB-MEN, ‘bias due to missing evidence’ refers to bias arising when some study results are unavailable because of their results. This may be, for example, because of large p-values, small magnitudes of effect, or harmful treatment effects. Such bias can be due to two types of missing evidence: i) the selective reporting of outcome results within studies published or otherwise known to exist, called *known unknowns* bias in the tool; ii) studies that remain entirely unpublished and are not known to exist, referred to as *unknown unknowns* bias.

In NMA, estimates of treatment effects are derived by combining direct and indirect evidence. Direct evidence refers to evidence about pairs of treatments that have been directly compared within studies. Indirect evidence refers to evidence on pairs of treatments that is “indirectly” derived from the sources of direct evidence via a common comparator or chain of comparisons (see also Box 1). In ROB-MEN, we first evaluate the likely risk of bias due to missing evidence for each possible pairwise comparison between the interventions of interest, irrespective of the availability of direct evidence. We then assess the impact of each pairwise comparison on the NMA by considering its percentage contribution to each NMA estimate. The relative treatment effects in an NMA are estimated using both direct and indirect evidence (‘mixed’ estimates), only direct evidence (‘only direct’ estimates) or only indirect evidence (‘only indirect’ estimates) depending on which comparisons are investigated in the identified studies (see also Glossary, Box 2).

At the core of the tool are two tables that record the various assessments for each pairwise comparison and each NMA estimate:

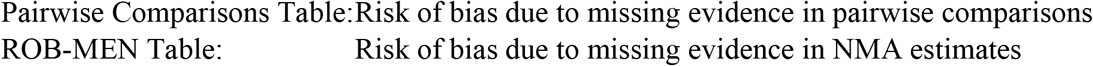

Both tables are completed separately for each outcome, i.e. for each NMA in the review. The Pairwise Comparisons Table facilitates the assessments in the ROB-MEN Table. The assessments in the Pairwise Comparisons Table largely follow the standard ROB-ME tool for pairwise meta-analysis [9]. Like ROB-ME, we consider not only the studies contributing to the current NMA but also the studies contributing to NMAs of any other outcomes in the systematic review. Such studies are informative about the possibility of selective non-reporting of the outcome being addressed in the current NMA. What is different about the ROB-MENm tool is that we need to consider all possible pairwise comparisons that could be made among the interventions in the network. This is because there may be missing evidence on any of the direct comparisons that *were* observed among the included studies, and also missing evidence on any of the comparisons that were *not* observed among the included studies. The output of the Pairwise Comparisons Table is a judgement about whether there is concern about bias due to missing evidence for each of the possible comparisons made from the interventions in the network.

The ROB-MEN Table is the main output of interest from the tool. It combines the outputs from the Pairwise Comparisons Table with (i) information about the structure and the amount of data in the network and (ii) the potential impact of missing evidence on the NMA results, to reach a judgement about risk of bias for each NMA estimate. The structure and amount of data in the network are represented by the percentage contributions of each piece of direct evidence to each NMA estimate. NMA estimates will be at higher risk of bias if they have high contributions from direct evidence considered to be susceptible to bias. We use network meta-regression methods targeting small-study effects to assess the potential impact of reporting bias on the results.

To fill in both the Pairwise Comparison Table and the ROB-MEN Table, we have developed an R Shiny web application (https://cinema.ispm.unibe.ch/rob-men/) that automates many of the steps required by the ROB-MEN process, as described in Box 3 and Box 4.

#### Box 3

**Instructions for filling in the Pairwise Comparisons Table**

**Box 3: Instructions for filling in the Pairwise Comparisons Table**
| Task | Implementation in R Shiny web application |
| --- | --- |
| List all possible pairwise comparisons between the interventions involved in the network and organize them in three groups “observed for this outcome”, “observed for other outcomes”, “unobserved”. | Automated |
| Enter in column 1 the number of studies (and total number of participants randomized in brackets) reporting the outcome of interest for the comparison and in column 2 the total number of studies identified for the comparison (and the relevant total number of participants randomized in brackets). Enter 0 in column 1 for comparisons “observed for other outcomes” and “unobserved” and in column 2 for “unobserved” comparisons. | Automated |
| Assess the level of risk for the “known unknowns” (selective outcome reporting) using a classification system and enter in column 3 “NA”, “undetected bias”, or “suspected bias favouring treatment X” according to which treatment is believed to be favoured. | Manual/<br>automated |
| Assess the level of bias due to “unknown unknowns” (publication bias) and enter in column 4 “undetected bias” or “suspected bias favouring treatment X” according to which treatment is believed to be favoured. | Manual |
| Merge “qualitatively” the assessments for the “unknown unknown” bias and the “known unknown” bias, as applicable, following the algorithm in Figure 1. For each comparison, enter in column 5 either “undetected bias” or “suspected bias favouring treatment X” according to the treatment favoured. | Automated/ manual |

#### Box 4

**Instructions for filling in the ROB-MEN Table**

**Box 4: Instructions for filling in the ROB-MEN Table**
| **Task** | **Implementation in R Shiny web application** |
| --- | --- |
| List all network estimates and organize them into two groups, “mixed/only direct” and “only indirect”. | Automated |
| Enter in column 1 the percentage contribution of direct evidence with suspected bias favouring the first treatment, in column 2 the percentage contribution of direct evidence favouring the second treatment, and in column 3 the total percentage contribution of direct evidence with suspected bias. | Automated |
| Evaluate the contribution from comparisons with suspected bias to each estimate and enter in column 4 “No substantial contribution from bias”, “Substantial contribution from bias balanced” or “Substantial contribution from bias favouring X” according to the treatment favoured. | Manual |
| Copy the final judgements (“undetected bias” or “suspected bias favouring treatment X” according to the treatment favoured) from column 6 of the Pairwise Comparisons Table to column 5 of the ROB-MEN Table only for comparisons with indirect evidence. | Automated |
| Run a network meta-regression model for small-study effects and enter the NMA estimates adjusted for the most precise study in column 7, alongside the relative NMA summary effect in column 6. | Automated |
| Evaluate the presence or absence of small-study effects and enter in column 8 “No evidence of small-study effects” or “Small-study effects favouring treatment X” according to the treatment favoured by the small studies. | Manual |
| For each NMA estimate enter in column 9 “high risk”, “some concerns” or “low risk” according to the algorithm rules in Box 5. | Automated |

### 2.2 Risk of bias due to missing evidence in pairwise comparisons (Pairwise Comparisons Table)

This section describes in more detail the steps required for assessing bias due to missing evidence in all possible pairwise comparisons. Each description is followed by a short instruction for filling in the relevant column in the Pairwise Comparisons Table. A summary of the process is provided in Box 3. The steps are illustrated using the network of non-invasive diagnostic modalities introduced in section 1 and Box 1 and the resulting Pairwise Comparison Table is given in Table 2.

#### 2.2.1 List of the pairwise comparisons

Once the studies have been identified for each outcome included in the review, users list all possible pairwise comparisons between the interventions involved in the network, that is, all combinations of two treatments. These constitute the rows of the table for assessing the risk of bias due to missing evidence for the pairwise comparisons (Pairwise Comparisons Table) for a specific outcome. We organise the comparisons into three groups as follows:

A. “observed for this outcome”: the comparisons for which there is direct evidence contributing to the NMA for the current outcome
B. “observed for other outcomes”: the pairwise comparisons for which there is direct evidence only for other outcomes in the systematic review
C. “unobserved”: the pairwise comparisons that have not been investigated in any of the identified studies in the systematic review

### Application to illustrative example

Of the possible 15 comparisons, 8 were observed for the outcome of interest (group A) and the remaining 7 were all unobserved (group C) i.e. there was no comparison observed only for other outcomes (group B).

#### 2.2.2 Number of studies and participants randomized in observed comparisons reporting outcome of interest or other outcomes (columns 1 and 2)

In the Pairwise Comparisons Table, we first list the number of studies that report results for the current outcome for the corresponding pairwise comparison. This will be non-zero for comparisons “observed for this outcome” (group A), and zero for “observed for other outcomes” (group B) and “unobserved” (group C) groups. We add in brackets the total sample size by adding up all participants randomized in the studies investigating the specific comparison for that outcome. Then, we enter the total number of studies identified in the systematic review making the corresponding comparison, again adding in brackets the total sample size for all studies examining that specific comparison for any outcome. By definition, the comparisons “observed for other outcomes” will have zero in the first column, while the “unobserved” comparisons will have zero in both columns.

#### 2.2.3 Evaluate the “known unknowns” bias (column 3; possible bias levels: “NA”, “undetected bias”, “suspected bias favouring X”)

Evaluation of bias due to selective non-reporting of results takes place for studies identified in the review but missing from the synthesis because results known (or presumed) to have been generated are unavailable. This bias is associated with studies reporting other outcomes but not the outcome of interest. The studies need to be evaluated for selective non-reporting of results. This could be done using study-specific tools such as the Outcome Reporting Bias In Trials (ORBIT) [19] or its simplified version described in Step 2 of the ROB-ME tool [9]. Then, the likely impact of the missing results across all studies may be assessed using the signalling questions below to reach an overall judgement of “undetected bias” or “suspected bias favouring X” for each comparison, as reported in Table 1.

**Table 1:** Responses to signalling questions to reach an overall judgement for the “known unknowns” of comparisons “observed for this outcome” or “observed for other outcomes”.

| Signalling question | Responses for each comparison (group A and B only) |  |  |
| --- | --- | --- | --- |
| 1 | Yes | Yes | No |
| 2 | Yes | No | - |
| Overall judgment | *Suspected bias (favouring X)* | *Undetected bias* | *Undetected bias* |

The signalling questions are the following:

1. Was there any eligible study for which results for the outcome of interest were unavailable, likely because of the P value, magnitude or direction of the result generated? (*Yes/No*)
2. (If Yes to previous question) Was the amount of information omitted from the synthesis sufficient to have a notable effect on the magnitude of the synthesized result? (*Yes/No*)

A thorough assessment of the “known unknowns” bias is likely to be labour intensive, but also very valuable as the impact of selective non-reporting or under-reporting of results can be quantified more easily than the impact of selective non-publication of an unknown number of studies [10]. However, for comparisons “observed for this outcome” if the number of studies (or the sample size) not reporting the outcome of interest (i.e. the difference between the numbers in column 2 and column 1) is small in comparison with the number of studies (or the total sample size) reporting the outcome (column 1), the final judgement from the assessment of these few studies may not be very informative and not affect the “known unknowns” judgement. In this case, reviewers might decide not to carry out the assessment above and assign “undetected bias” to the relevant comparison. “Undetected bias” is also assigned in the situation that no study is suspected of selective non-reporting or under-reporting of results for a specific comparison (i.e. the numbers in the first two columns are equal). For all “unobserved” comparisons (group C) a level of “NA” is assigned because the assessment is not applicable.

### Application to illustrative example

Other than those included in the analysis, there did not seem to be any extra studies identified in the review which did not report results for the outcome of interest for the comparisons “observed for this outcome”. Therefore, we can assume that there is no selective outcome reporting bias for this example and we assign “undetected bias” for the “known unknowns” to all comparisons in this group. Comparisons in “unobserved” group (group C) are assigned “NA” level as they cannot be judged for selective outcome reporting bias. See column 3 of Table 2 for the “known unknowns” judgements for all comparisons.

**Table 2:** Pairwise Comparisons Table for the network of non-invasive diagnostic modalities for detection of coronary artery disease in patients with low risk acute coronary syndrome. CCTA: coronary computed tomographic angiography; CMR: cardiovascular magnetic resonance; ECG: electrocardiogram; Echo: echocardiography; SPECT-MPI: single photon emission computed tomography-myocardial perfusion imaging; SR: systematic review.

|  | 1 | 2 | 3 | 4 | 5 |
| --- | --- | --- | --- | --- | --- |
| Pairwise comparisons | Number of studies in each comparison |  | “known unknowns” | “unknown unknowns” | Overall bias |
|  | Reporting this outcome (sample size) | Total identified in the SR (total sample size) | Classification system & signalling questions | Qualitative signals & quantitative considerations | Synthesizing judgements |
| **Group A:**<br>**observed for this outcome** |  |  |  |  |  |
| CCTA vs Exercise ECG | 1 (562) | 1 (562) | Undetected bias | Undetected bias | Undetected bias |
| CCTA vs SPECT-MPI | 2 (1149) | 2 (1149) | Undetected bias | Suspected bias favouring CCTA | Suspected bias favouring CCTA |
| CCTA vs Standard care | 7 (4015) | 7 (4015) | Undetected bias | Suspected bias favouring CCTA | Suspected bias favouring CCTA |
| CMR vs Standard care | 2 (214) | 2 (214) | Undetected bias | Suspected bias favouring CMR | Suspected bias favouring CMR |
| Exercise ECG vs Standard care | 1 (130) | 1 (130) | Undetected bias | Undetected bias | Undetected bias |
| Exercise ECG vs Stress Echo | 4 (1086) | 4 (1086) | Undetected bias | Suspected bias favouring Stress Echo | Suspected bias favouring Stress Echo |
| SPECT-MPI vs Standard care | 2 (4165) | 2 (4165) | Undetected bias | Undetected bias | Undetected bias |
| Standard care vs Stress Echo | 1 (132) | 1 (132) | Undetected bias | Suspected bias favouring Stress Echo | Suspected bias favouring Stress Echo |
| **Group B:**<br>**observed for other outcomes** |  |  |  |  |  |

| **Group C:**<br>**Unobserved** |  |  |  |  |  |
| --- | --- | --- | --- | --- | --- |
| CCTA vs<br>CMR | 0 | 0 | NA | Undetected bias | Undetected bias |
| CCTA vs<br>Stress Echo | 0 | 0 | NA | Suspected bias<br>favouring CCTA | Suspected bias<br>favouring CCTA |
| CMR vs<br>Exercise ECG | 0 | 0 | NA | Undetected bias | Undetected bias |
| CMR vs<br>SPECT-MPI | 0 | 0 | NA | Undetected bias | Undetected bias |
| CMR vs<br>Stress Echo | 0 | 0 | NA | Undetected bias | Undetected bias |
| Exercise ECG vs<br>SPECT-MPI | 0 | 0 | NA | Suspected bias<br>favouring SPECT-MPI | Suspected bias<br>favouring SPECT-MPI |
| SPECT-MPI vs<br>Stress Echo | 0 | 0 | NA | Suspected bias<br>favouring SPECT-MPI | Suspected bias<br>favouring SPECT-MPI |

#### 2.2.4 Decide the “unknown unknowns” bias (column 4, possible bias levels: “undetected bias”, “suspected bias favouring X”)

This refers to studies undertaken but not published, so review authors are unaware of them. Each comparison is assessed for risk of bias using primarily qualitative and secondarily quantitative considerations, if applicable.

A qualitative judgement is made for all comparisons to assign a level of *undetected* or *suspected bias*. Conditions that may indicate *suspected bias* include but are not limited to: a failure to include unpublished data and data from grey literature; the meta-analysis is based on a small number of positive early findings, for example for a drug newly introduced on the market (as early evidence is likely to overestimate its efficacy and safety); previous evidence documenting the presence of publication bias for that specific comparison. Whereas conditions suggesting *undetected bias* may include: data from unpublished studies have been identified, and their findings agree with those in published studies; there is a tradition of prospective trial registration in the field.

For comparisons with at least 10 studies (in column 1) judgements can additionally consider statistical techniques such as contour-enhanced funnel plots, which can indicate whether results appear to have been suppressed because they did not reach statistical significance [3], appropriate regression models and associated statistical tests for small-study effects [1,5,6,20– 22], and selection model for pairwise meta-analysis (e.g. Copas [7]). With any of these approaches, the direction of any suspected bias should be noted: the bias will generally be in favour of the treatment favoured most in the smaller studies.

### Application to illustrative example

None of the observed direct comparisons had 10 or more studies available and were therefore not eligible for the “unknown unknowns” bias assessment using graphical and statistical methods. Using the qualitative signals for the “unknown unknowns”, we considered CCTA vs SPECT-MPI, CCTA vs standard care, and CCTA vs stress echo to be at suspected bias favouring CCTA because the latter is a new non-invasive easily-accessible imaging technology so we assumed that any unpublished study involving this intervention reported unfavourable results for the investigators. We also considered CMR vs standard care to be at suspected bias favouring CMR, for similar reasons. We suspected exercise ECG vs stress echo and standard care vs stress echo to be biased in favour of stress echo as this is a more contemporary method with higher diagnostic accuracy. Finally, we judged exercise ECG vs SPECT-MPI and SPECT-MPI vs stress echo to be at suspected bias in favour of SPECT-MPI because this was the first widely available non-invasive imaging technology for functional assessment of the heart and was considered the gold-standard method for several years, especially in the US, without any strong evidence of clinical benefit over other methods. We assigned “Undetected bias” to all other comparisons. See column 4 of Table 2 for the “unknown unknowns” judgements for all comparisons.

#### 2.2.5 Overall risk of bias for pairwise comparisons (column 5; possible bias levels: “undetected bias”, “suspected bias favouring X”)

The last step in the Pairwise Comparisons Table is to combine the levels of risk assigned in the previous steps into a final judgement. This is also described in the flowchart in Figure 1. ***For the unobserved comparisons (group C)*** this will be the same as the judgement made for the “unknown unknown” bias, as this is the only assessment applicable to these comparisons. ***For the comparisons observed for other outcomes (group B)*** the overall judgement will consider qualitative assessments for both the “known unknown” and the “unknown unknown” bias. The assessment of selective outcome reporting bias (“known unknowns”) is likely to be the most valuable because its impact can be quantified more easily than that of publication bias (“unknown unknowns”). Therefore, if the reviewer deems a comparison to be at suspected bias due to selective outcome reporting, then the final judgement should be that the comparison has suspected bias regardless of the findings in the “unknown unknown” assessment.

**Figure 1:**
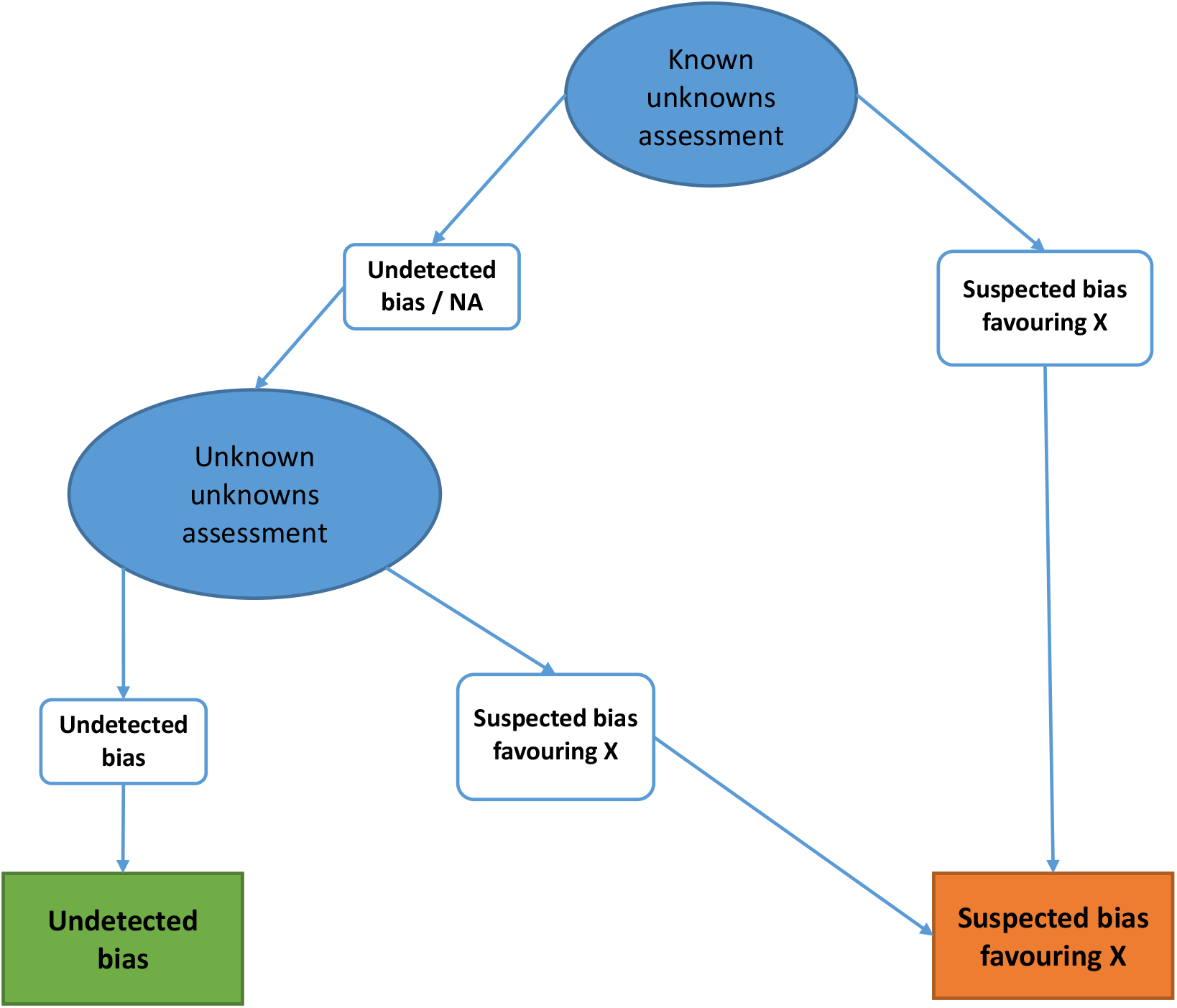
Algorithm for assessing overall risk of bias due to missing evidence in pairwise comparisons.

***The overall judgement for comparisons observed for this outcome (group A)*** will follow the same recommendations in the previous paragraph, with the only difference that graphical and statistical methods could also be included for the “unknown unknowns” assessment. The latter can be useful in cases where it is difficult to assess selective outcome reporting reliably e.g. when the search for studies is not comprehensive and/or the protocol and records from trial registries were unavailable. Therefore, in such cases, if the quantitative methods indicate evidence of publication bias, then the reviewer should consider that comparison to be with suspected bias.

### Application to illustrative example

Following the algorithm described above, we merge the previous assessments into an overall bias for pairwise comparisons and report it in the last column of the Pairwise Comparison Table (Table 2). Since there was no selective outcome reporting bias (“known unknowns”) assessment, the overall bias for comparisons “observed for this outcome” will only consider the “unknown unknowns” assessment. Therefore, we judged CCTA vs SPECT-MPI, CCTA vs standard care and CMR vs standard care to be at suspected bias favouring the first treatment, respectively; CCTA vs stress echo and exercise ECG vs stress echo to be at suspected bias favouring stress echo. Also, for “unobserved” comparisons the only available assessment is the one for “unknown unknowns” bias so the relevant judgment will constitute also the final judgement. In this case, we suspected CCTA vs stress echo, exercise ECG vs SPECT-MPI, and SPECT-MPI vs stress echo to be at suspected bias favouring CCTA and SPECT-MPI, respectively.

### 2.3 Risk of bias due to missing evidence in NMA estimates (ROB-MEN Table)

Once the assessments of overall bias for each pairwise comparison are complete, we integrate them in the assessment of risk of bias for each NMA estimate. This is achieved by combining the contribution of the comparisons to the network estimate with the additional risk of bias for indirect comparisons (because of missing direct evidence) and any evidence of small-study effects. We consider all NMA estimates and list them as rows of the ROB-MEN Table. We organize the estimates into two groups, “mixed/only direct” and “only indirect”, depending on the type of evidence contributing to each estimate (see also Glossary, Box 2).

We describe here the detailed steps for filling in the relevant column in the ROB-MEN Table. A summary of the process is provided in Box 4. As for the risk of bias due to missing evidence in pairwise comparisons, we illustrate the steps by filling in the ROB-MEN Table for the network of non-invasive diagnostic modalities (Table 3).

**Table 3:**
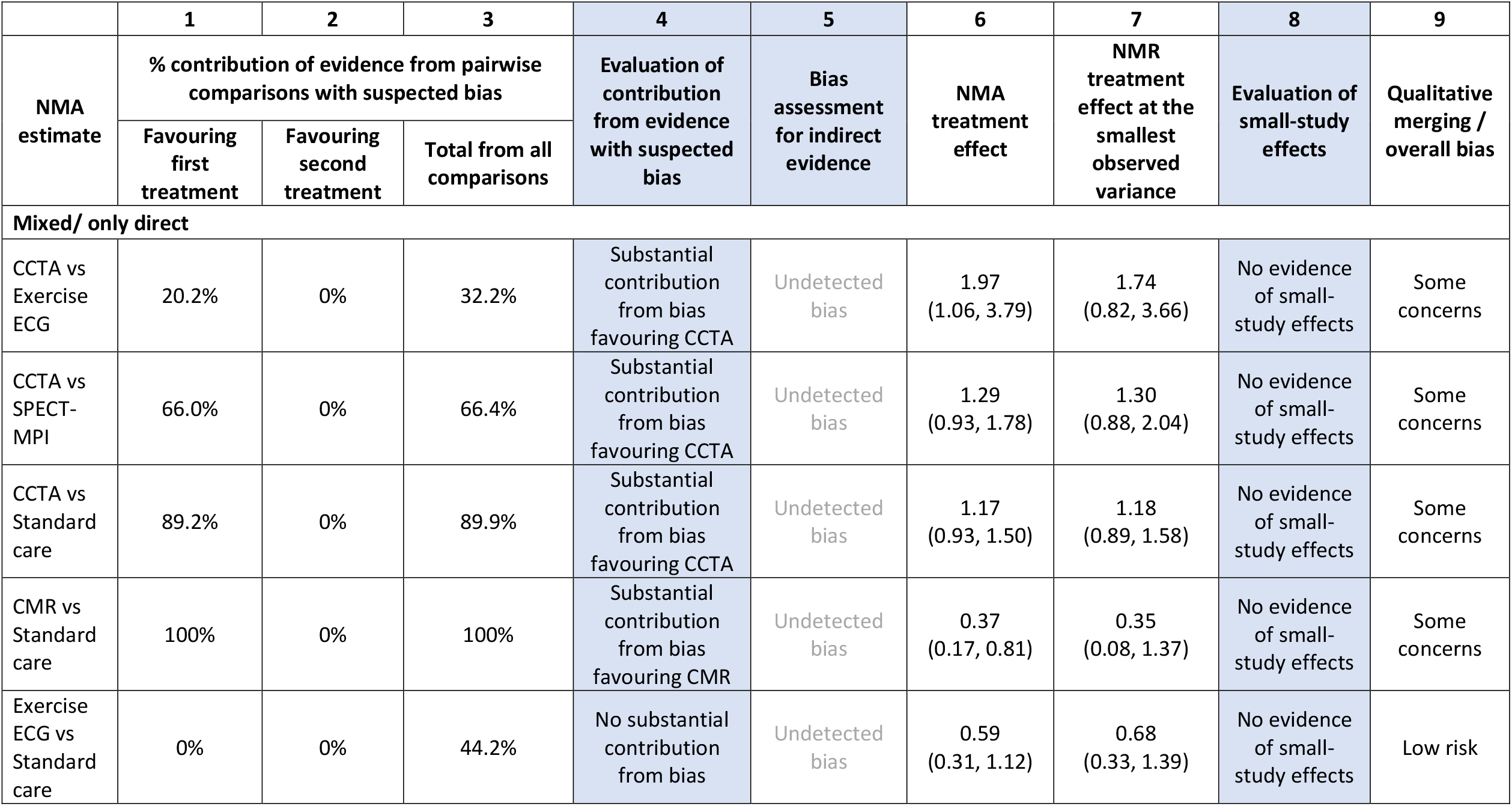
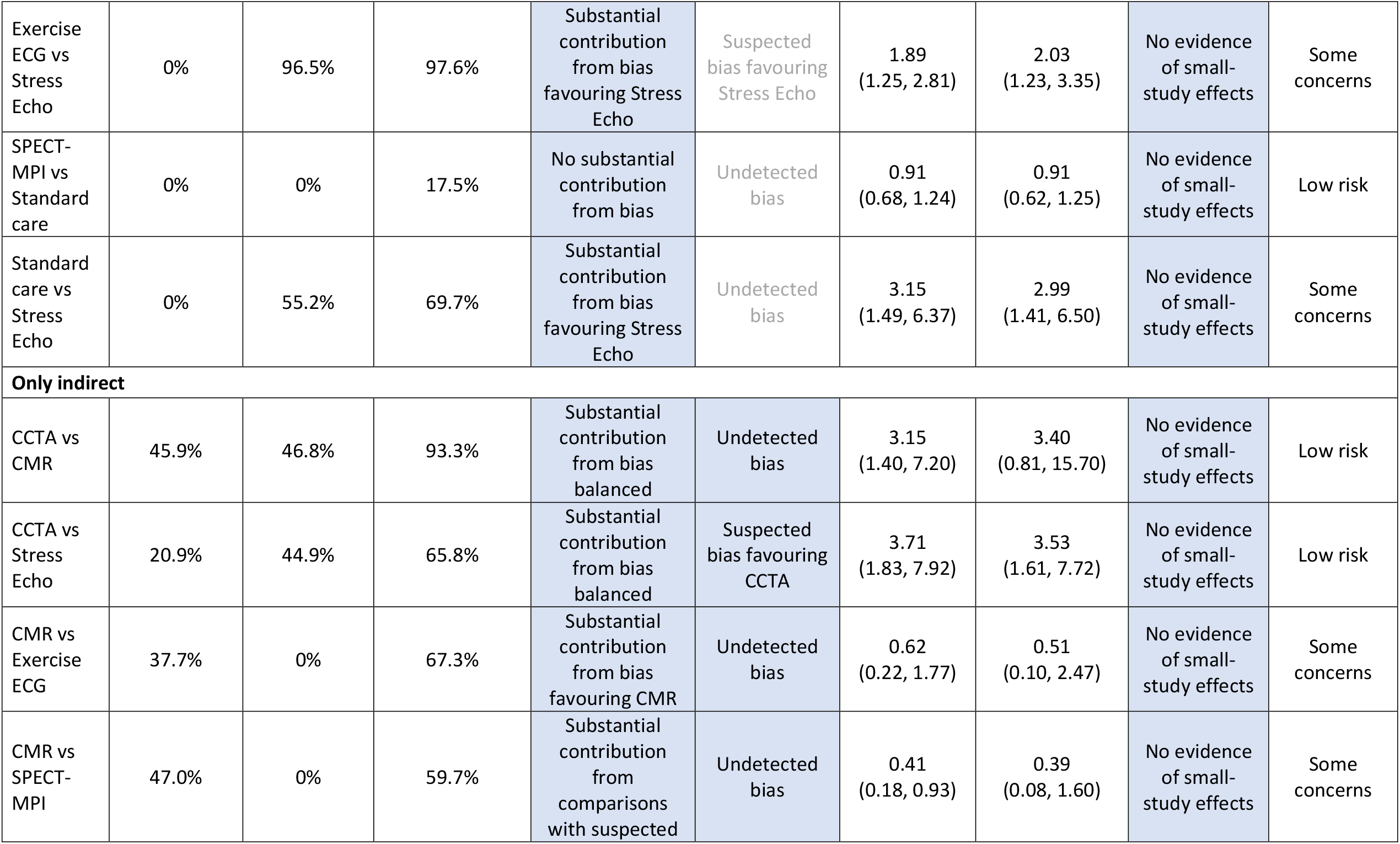
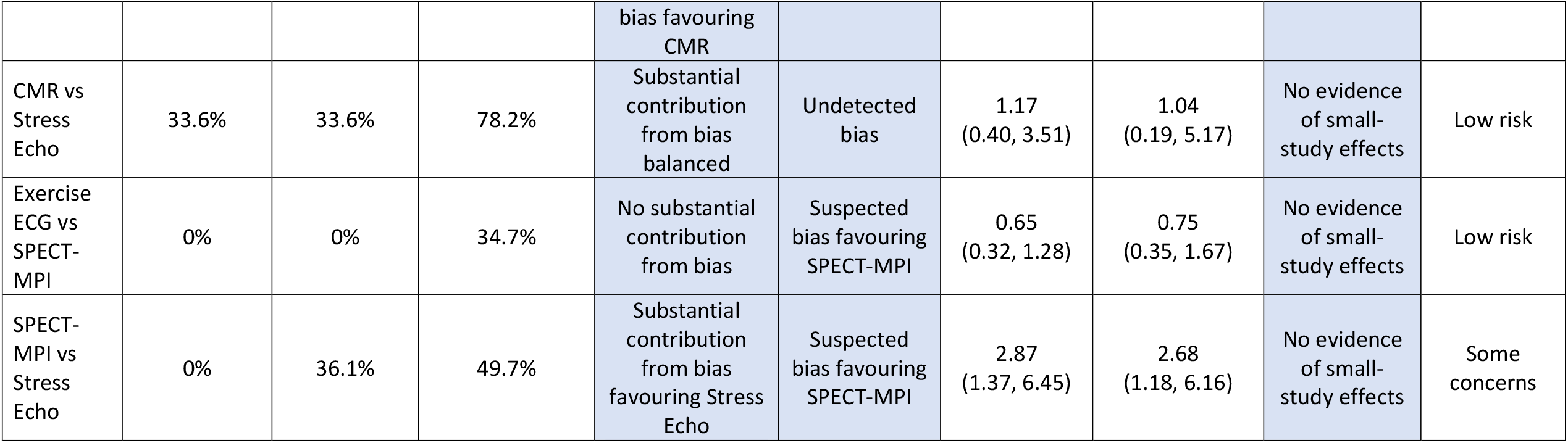
ROB-MEN Table for the network of non-invasive diagnostic modalities for detection of coronary artery disease in patients with low risk acute coronary syndrome. CCTA: coronary computed tomographic angiography; CMR: cardiovascular magnetic resonance; ECG: electrocardiogram; Echo: echocardiography; NMA: network meta-analysis; NMR: network meta-regression; SPECT-MPI: single photon emission computed tomography-myocardial perfusion imaging. Effects in column 6 and 7 are odds ratios and 95% credible intervals.

#### 2.3.1 Contribution of comparisons with suspected bias to the NMA estimates (columns 1, 2, 3, 4; possible levels: “No substantial contribution from bias”, “Substantial contribution from bias favouring X”, “Substantial contribution from bias balanced”)

The first step in the assessment of bias due to missing evidence in an NMA estimate is to consider the contribution matrix of the network. This matrix has the NMA relative treatment effect estimates as rows and the sources of direct evidence (i.e. the comparisons “observed for this outcome”, group A) as columns. Each cell entry provides the percentage contribution that each comparison with direct evidence makes to the calculation of the corresponding NMA relative treatment effect [23].

We focus on the direct evidence with suspected risk of bias from the overall bias assessment from the Pairwise Comparisons Table. We consider any specific percentage contribution from direct evidence with suspected bias favouring either one of the two treatments in each estimate and enter these in the first and second column, respectively. Additionally, we add up the total percentage contribution any direct evidence with suspected bias makes to each NMA relative effect, regardless of the direction and treatments involved, and report this in the third column of the ROB-MEN Table for descriptive purposes only.

Finally, the results of the evaluation of the contribution from comparisons with suspected bias is reported in the fourth column. This is represented by one of the levels according to whether there is substantial contribution favouring either one of the treatments or if the contribution is split more or less equally between evidence with bias in the opposite direction. Specifically:

- *No substantial contribution from bias*: there is no substantial contribution from evidence at suspected bias favouring either one of the two treatments;
- *Substantial contribution from bias balanced*: there is substantial contribution from evidence at suspected bias but it is split more or less equally between evidence with bias favouring one of the treatments and evidence with bias favouring the other treatment;
- *Substantial contribution from bias favouring X*: there is substantial contribution from evidence at suspected bias favouring one of the two treatments (say X).

### Application to illustrative example

We consider the network percentage contribution matrix (Appendix Table 1) to calculate the contributions from the five comparisons with direct evidence (“observed for this outcome”) with suspected bias. For each NMA estimates we enter in the third column of the ROB-MEN Table (Table 3) the total contribution from the five sources of direct evidence judged at suspected bias, regardless of its direction; then, where applicable, we separate the total contribution from these sources at suspected bias favouring the first treatment of the estimate and the total contribution from those favouring the second treatment in the estimate. Among the mixed estimates, six of them have a clear separation of high contribution coming from biased evidence between the two treatments, like CCTA vs exercise ECG, CCTA vs SPECT-MPI, CCTA vs standard care, CMR vs standard care, exercise ECG vs stress echo and standard care vs stress echo. Among the indirect estimates, only three estimates showed such clear separation (CMR vs exercise ECG, CMR vs SPECT-MPI and SPECT-MPI vs stress echo) while in other three (CCTA vs CMR, CCTA vs stress echo, CMR vs stress echo) the percentage contribution is split between sources of evidence at suspected bias in the opposite direction. The relevant level for this step is entered in column 4 of the ROB-MEN Table (Table 3)

#### 2.3.2 Additional risk of bias for indirect estimates (column 5; possible levels: “undetected bias”, “suspected bias favouring X”)

Indirect relative effects are calculated from sources of direct evidence in the Pairwise Comparisons Table with contributions as shown in the contribution matrix. However, the absence of direct evidence for these indirect comparisons will lead to bias if studies that actually made the direct comparison are missing for reasons associated with their results.

Therefore, for the indirect estimates we need to account for this potential source of bias, which is represented by the final judgement of the overall bias from the Pairwise Comparisons Table.

### Application to illustrative example

We copy the final judgements from column 5 of the Pairwise Comparisons Table (Table 2) into column 5 of the ROB-MEN Table (Table 3). Even though the full column is copied, this additional source of bias is only considered for the indirect estimates. Among these, three (CCTA vs stress echo, exercise ECG vs SPECT-MPI, SPECT-MPI vs stress echo) were at suspected bias favouring CCTA and SPECT-MPI, respectively.

#### 2.3.3 Evaluate small-study effects in NMA (columns 6, 7, 8; possible levels: “No evidence of small-study effects”, “Small-study effects favouring X”)

To evaluate small-study effects, we run a network meta-regression model (NMR) with a measure of precision (e.g. variance or standard error) as covariate. We use this model to generate an adjusted relative effect, by extrapolating the regression line to the smallest observed variance (the ‘largest’ study) independently for each comparison. To assess the presence of small-study effects we compare the obtained adjusted estimates with the original (unadjusted) estimates by looking at the overlap of their corresponding confidence (or credible) intervals. A lack of overlap between the two intervals (or between one estimate and the interval for the other estimate) is an indication that the small studies show different effects from the larger studies. We might be particularly concerned that the NMA effect is more in favour of the treatment favoured in small studies compared with the NMR effect. Note that this indication of small-study effects assumes there is no other explanation for the difference between the original and the adjusted estimates i.e. it is not explained by other covariates.

The result of the evaluation of small-study effects is reported in the penultimate column of the ROB-MEN Table as a judgement indicating whether there is evidence of small-study effects and, if so, which treatment is favoured by the small studies.

### Application to illustrative example

We run a NMR model using the variance of the estimate (pooled variance for multi-arm studies) as a covariate to investigate small-study effects in the whole network. The adjusted estimates via extrapolation to the smallest observed variance are reported in column 7 of the ROB-MEN Table (Table 3) next to the original NMA summary effect (column 6). None of the NMR estimates are markedly different from their unadjusted counterparts and there seem to be a good overlap of the two credible intervals for all estimates. Therefore, “No evidence of small-study effects” is reported in column 8 for all the estimates.

#### 2.3.4 Overall risk of bias for NMA estimates (column 9; possible bias levels: “low risk”, “some concerns”, “high risk”)

The algorithm rules for assigning a final judgement on the overall risk of bias due to missing evidence for NMA estimates are described in Box 5. This should consider the contribution from comparisons with suspected bias (column 4) and any substantial difference between the original and NMA effects adjusted for the most precise study (column 8). For NMA indirect estimates, the conclusions for overall bias of comparisons in column 5 should also be considered in the final judgement.

If there is substantial contribution from evidence with suspected bias, we have concerns regarding the risk of bias for that estimate. However, if this contribution is split more or less equally between evidence with bias favouring one of the treatments and evidence with bias favouring the other treatment, then we might hypothesize the two biases in the opposite direction cancel out, under the assumption that the magnitude of the bias is roughly the same in the two directions. Concerns about the risk of bias are then defined by the overall bias of unobserved comparisons (for NMA indirect estimates) and the evidence about small-study effects.

### Application to illustrative example

Given that most of the mixed estimates have substantial contribution from biased evidence favouring one of the two treatments but there was no evidence of small-study effects for any of the estimates, we have some concerns about the risk of bias due to missing evidence except for exercise ECG vs standard care and SPECT-MPI vs standard care where the level was decreased to “Low risk” due to lack of substantial contribution from biased evidence favouring either one of the two treatments. Similarly, we assigned a level of “Some concerns” to some of the indirect estimates, where the substantial contribution from biased evidence was favouring either one of the two treatments (CMR vs Exercise ECG, CMR vs SPECT-MPI, SPECT-MPI vs Stress Echo). All the other indirect estimates were assigned a level of “Low risk” of bias due to missing evidence because the substantial contribution from evidence at suspected bias was either absent or split equally between sources of evidence with bias in the opposite direction, there was no additional bias coming from the indirect comparison assessed in the Pairwise Comparisons Table and no evidence of small-study effects. No estimate was judged to be at high risk of bias due to missing evidence.

Our final judgements for the overall risk of bias due to missing evidence in the network are reported in column 9 of the ROB-MEN Table (Table 3) as follows:

- no NMA estimates at high risk of bias due to missing evidence;
- six NMA estimates at low risk of bias due to missing evidence (exercise ECG vs standard care, SPECT-MPI vs standard care, CCTA vs CMR, CCTA vs stress echo, CMR vs stress echo, exercise ECG vs SPECT-MPI);
- the remaining NMA estimates with some concerns about bias due to missing evidence.

#### Box 5

**Algorithm rules for assigning final judgement on the overall risk of bias due to missing evidence for NMA estimates**

|  |  |
| --- | --- |
| Low risk | There is no substantial contribution from evidence with suspected bias favouring one of the two treatments,<br>OR<br>There is substantial contribution from evidence at suspected bias but it is split more or less equally between evidence with bias favouring one of the treatments and evidence with bias favouring the other treatment<br>AND<br>There is no evidence of small-study effects favouring one of the two treatments<br>OR<br>[For indirect estimates only] There is no suspected bias favouring one of the two treatments from the assessment of indirect evidence. |
| Some concerns | All other combinations |
| High risk | There is substantial contribution from evidence with suspected bias favouring one of the two treatments, say X<br>AND<br>There is evidence of small-study effects favouring the same treatment X<br>OR<br>[For indirect estimates only] There is suspected bias favouring that treatment X from the assessment of indirect evidence. |

### 3 Application of ROB-MEN to a network comparing 18 antidepressants

We apply the ROB-MEN to assess the risk of bias due to missing evidence in a network of 18 antidepressants using only head-to-head studies (i.e. only studies investigating active interventions) from the review by Cipriani et al [17]. The outcome of interest is response to treatment defined as the number of patients who had a reduction of at least 50% on the total score between baseline and week 8 (range 4–12 weeks) on a standardized observer-rating scale for depression [24].

#### Pairwise Comparisons Table

There are 153 possible comparisons between the 18 drugs, 70 were reported for the outcome response (group A) and 2 comparisons (amitriptyline versus bupropion and amitriptyline versus nefazodone) were reported for other outcomes (dropouts and remission, group B). The remaining 82 comparisons were not investigated in any of the identified studies (“unobserved”, group C) and they are listed at the end of the table (Appendix Table 2).

The Pairwise Comparison Table starts with the “known unknowns” assessment. We carried this out only for the two comparisons in the “observed for other outcomes” group, both of them judged with undetected bias, and for those comparisons in the group “observed for this outcome” for which extra studies were identified that did not report the outcome of interest. We judged four of these to be at suspected bias because the extra studies did not fully report the results and were sponsored by the company manufacturing the drug favoured by the bias. We judged the other four comparisons as “Undetected bias” because we deemed the unavailable results unlikely to be missing due to unfavourable p-values or directions of the results generated, or because they were unlikely to affect the synthesized result notably. For example, the extra study in the comparison of bupropion versus paroxetine focused on suicidal ideation only and removed the relative items from the full depression score which, therefore, could not be included in the NMA. Another example is the extra study of fluoxetine versus paroxetine which, despite being suspected of selective outcome reporting bias, is unlikely to have a notable effect on the synthesized result given its small sample size (21 participants) relative to the large total sample size for the included studies (1364 participants). We assigned all the other direct comparisons “observed for this outcome” a level of “Undetected bias” in this step, while the assessment is not applicable for the 82 “unobserved” comparisons.

The “unknown unknowns” assessment could be carried out for all comparisons and the following logic was followed to reach a judgement. We considered that bias, when suspected, would favour the newest drug, according to the novel agent bias principle. The exceptions were comparisons involving agomelatine, paroxetine, bupropion and vortioxetine as the newest drug because the authors were able to obtain all the unpublished data from the manufacturers of these drugs. This qualitative consideration took priority also over our findings from contour-enhanced funnel plots and regression-based tests for small-study effects for those comparisons with at least 10 studies. In fact, based on the findings from these statistical techniques, neither amitriptyline versus fluoxetine nor citalopram versus escitalopram would be judged at suspected bias. However, we agreed our “unknown unknowns” judgement for both comparisons as “Suspected bias favouring the newest drug” because the review authors could not exclude the possibility of hidden studies with unfavourable results towards the newer drug in the comparison (fluoxetine and escitalopram).

Following the algorithm (Figure 1) to reach the overall bias judgement for pairwise comparisons, most of them were considered at “Suspected bias favouring the newest drug”. The only ones judged with undetected bias were the comparisons involving agomelatine and vortioxetine, as well as amitriptyline versus paroxetine, bupropion versus fluoxetine, bupropion versus paroxetine clomipramine versus paroxetine, fluoxetine versus paroxetine, fluvoxamine versus paroxetine, paroxetine versus sertraline, paroxetine versus trazodone, amitriptyline versus bupropion, amitriptyline versus clomipramine, bupropion versus clomipramine, and citalopram versus paroxetine. The judgements for all pairwise comparisons are reported in the last column of the Pairwise Comparisons Table (Appendix Table 2).

### ROB-MEN Table

Once the Pairwise Comparison Table is complete with all judgements, we move to the ROB-MEN Table. First, the overall risk of bias judgements for comparisons with direct evidence are combined with the results from the contribution matrix to calculate for each NMA estimate the contribution coming from direct evidence at suspected bias favouring either of the two treatments, and in total. We considered an estimate to have substantial contribution from evidence at suspected bias favouring one of the two treatments in the contrast if the difference between the first and second column (contribution from evidence at suspected bias favouring first and favouring second treatment, respectively) was at least 15 (in percentage points).

The bias assessment for indirect evidence is only considered for the “only indirect” estimates and is copied from the last column of the Pairwise Comparison Table. This potential risk for “missing studies” is particularly important for the indirect estimates because it drives the bias evaluation to a “high risk” level in case there is also substantial contribution from direct evidence with suspected bias in the same direction.

The last part of the risk of bias assessment for the network estimate involves running a NMR model to evaluate the presence (or absence) of small-study effects. We run the model using the smallest observed variance as a covariate and assuming unrelated coefficients with a prespecified prior, *t*(0.*u*^2^.1) where *u* is again the largest maximum likelihood estimator in single trials. All NMA estimates and their adjusted counterpart were similar and their credible intervals had a good level of overlap, providing no evidence of small-study effects.

Following the algorithm rules set out in Box 4 we assign the final judgements on the overall risk of bias due to missing evidence to the NMA estimates and report it in the last column of the ROB-MEN Table (Appendix Table 3). Most estimates were judged with some concerns or at low risk of bias. In particular, none of the contrasts involving agomelatine, paroxetine, venlafaxine or vortioxetine were at high risk of bias.

All 153 NMA estimates with their relative ROB-MEN levels are reported in Table 4.

**Table 4:**
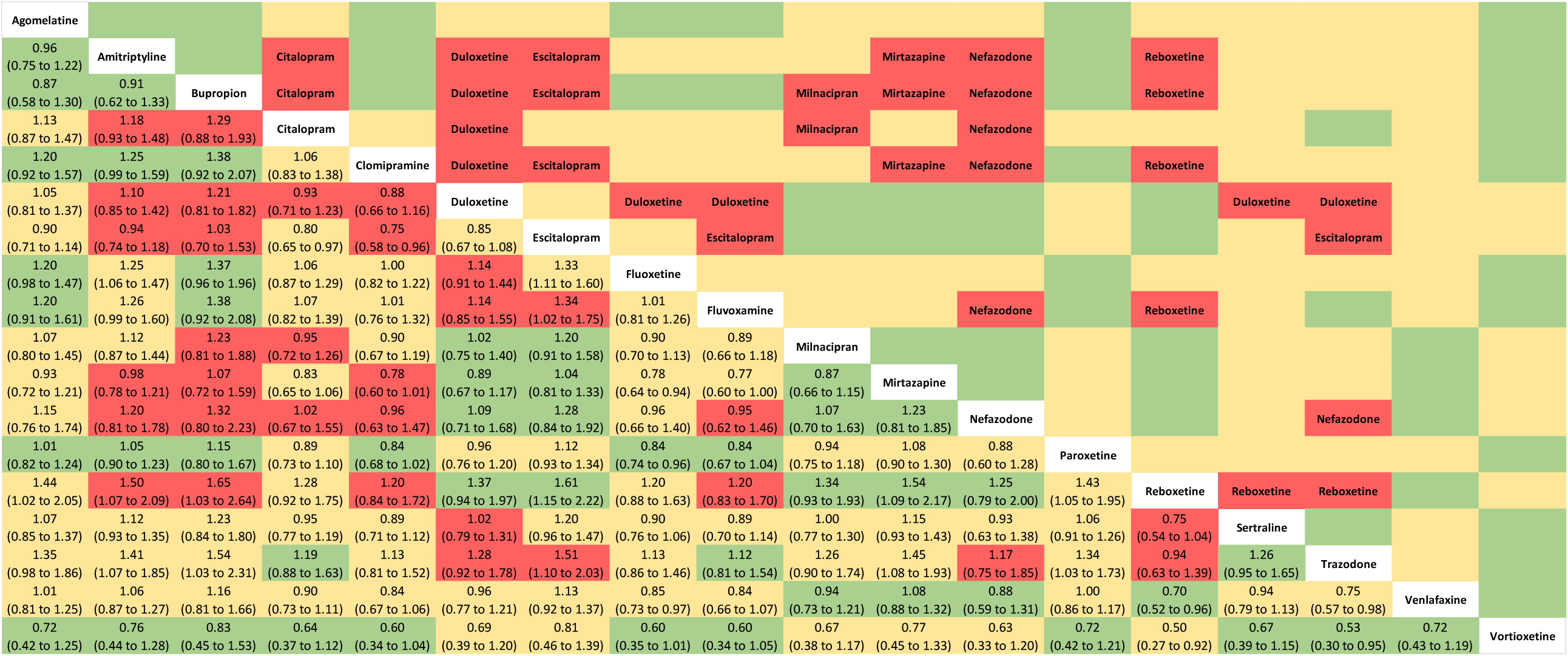
League table of the NMA estimated effects and corresponding risk of bias due to missing evidence for the network of 18 antidepressants. The values in the lower triangle represent the relative treatment effect (odds ratios and 95% credible intervals) of the treatment on the top (column) versus the treatment on the row. Colours indicate the ROB-MEN levels: green = Low risk; yellow: Some concerns; red = High risk. Names in the upper triangles indicates the treatment favoured by the bias in the high risk estimates (red cells). Risk of bias assessments are semi-automated in the ROB-MEN Shiny app.

## 4 Conclusion

To our knowledge, ROB-MEN is the first tool for assessing the risk of bias due to missing evidence in NMA. ROB-MEN builds on an approach recently proposed for pairwise meta-analysis [9,10] and integrates it into the NMA setting. Specifically, the assessments for selective outcome reporting and publication bias in pairwise comparisons are combined with (i) the percentage contribution of direct evidence for each pairwise comparison to the NMA estimates, (ii) evidence about the presence of small-study effects and (iii) any bias arising from unobserved comparisons.

We demonstrated with our examples that our tool is applicable to all NMAs, including very large and complex networks, for which the risk of bias assessment can be lengthy and labour-intensive. The R Shiny web application we have developed (https://cinema.ispm.unibe.ch/rob-men/) automates many of the ROB-MEN steps, therefore making the process much simpler and straightforward. The user uploads the raw data for their network of interventions, so that the app run the analysis required by the ROB-MEN. Once the user has evaluated the risk of bias for all pairwise comparisons and NMA estimates, the app produces the Pairwise Comparisons Table and ROB-MEN Table. As this project was also supported as part of updates to the CINeMA framework and software [25,26], we plan to incorporate the ROB-MEN tool within the reporting bias domain.

Our ROB-MEN methodology is not applicable in situations where there is an intervention disconnected from the network that is still of interest for decision-making, as it is not intended to cover comparisons involving such disconnected interventions. In case of disconnected networks, we recommend each subnetwork to be evaluated separately.

Like for any other evaluation of risk of bias or results’ credibility in evidence synthesis, many of the judgements in the ROB-MEN process involve subjective decisions of reviewers. Judging bias due to missing evidence is particularly challenging, particularly for publication bias, as reviewers often do not know whether studies were conducted and need to make informed guesses. However, the subjectivity of our approach, specifically in the pairwise comparisons step, is in line with the other existing techniques, as described in the Cochrane Handbook and ROB-ME tool [9,10]. Also, the novel and quantitative methods, such as the contribution matrix [23] and network meta-regression, that we integrated in the NMA estimate assessment, rely somewhat less on the reviewer’s subjectivity, achieving a balance between a pragmatic and rigorous approach. The tool will require studies for reliability and reproducibility of the assessments made by the users. When undertaking the ROB-MEN evaluation, we recommend reviewers to specify the criteria used and explain the reasoning behind the judgements to enhance transparency. We believe that ROB-MEN will help those performing NMA to reach better-informed conclusions and will greatly improve the toolbox of already available methods for evaluating the credibility of NMA results.

## Data Availability

Data sharing not applicable as no datasets generated and/or analysed for this study.

## Data availability statement

Data sharing not applicable as no datasets generated and/or analysed for this study.

