## Appendix for "Tool to assess risk of bias due to missing evidence in network meta-analysis (ROB-MEN): elaboration and examples"

Table 1: Contribution matrix for the network of non-invasive diagnostic modalities for detection of coronary artery disease in patients with low risk acute coronary syndrome. Cell values are the percentage contribution of the direct comparisons (columns) to the network estimates (rows).

|  | **CCTA vs Exercise ECG** | **CCTA vs**  **SPECT MPI** | **CCTA vs Standard care** | **CMR vs Standard care** | **Exercise ECG vs Standard care** | **Exercise ECG vs Stress Echo** | **SPECT MPI vs Standard care** | **Standard care vs Stress Echo** |
| --- | --- | --- | --- | --- | --- | --- | --- | --- |
| **CCTA vs CMR** | 0.81 | 5.41 | 40.52 | 46.75 | 0.53 | 0.28 | 5.41 | 0.28 |
| **CCTA vs Exercise ECG** | 51.64 | 2.02 | 18.16 | 0 | 14.2 | 5.98 | 2.02 | 5.98 |
| **CCTA vs SPECT-MPI** | 0.64 | 34.05 | 31.9 | 0 | 0.42 | 0.22 | 32.55 | 0.22 |
| **CCTA vs Standard care** | 1.17 | 8.12 | 81.04 | 0 | 0.8 | 0.37 | 8.12 | 0.37 |
| **CCTA vs Stress Echo** | 24.04 | 2.17 | 18.7 | 0 | 8.02 | 32.06 | 2.17 | 12.85 |
| **CMR vs Exercise ECG** | 15.63 | 2.04 | 13.58 | 37.65 | 15 | 7.03 | 2.04 | 7.03 |
| **CMR vs SPECT-MPI** | 0.13 | 5.88 | 5.74 | 46.97 | 0.08 | 0.05 | 41.09 | 0.05 |
| **CMR vs Standard care** | 0 | 0 | 0 | 100 | 0 | 0 | 0 | 0 |
| **CMR vs Stress Echo** | 10.96 | 1.51 | 9.45 | 33.64 | 9.27 | 20.23 | 1.51 | 13.41 |
| **Exercise ECG vs SPECT-MPI** | 20.83 | 12.99 | 7.84 | 0 | 14.83 | 6.95 | 29.62 | 6.95 |
| **Exercise ECG vs Standard care** | 23.1 | 2.72 | 20.38 | 0 | 29.99 | 10.54 | 2.72 | 10.54 |
| **Exercise ECG vs Stress Echo** | 1.14 | 0.15 | 0.99 | 0 | 1.09 | 94.26 | 0.15 | 2.23 |
| **SPECT-MPI vs Standard care** | 0.17 | 8.79 | 8.62 | 0 | 0.11 | 0.06 | 82.19 | 0.06 |
| **SPECT-MPI vs Stress Echo** | 13.6 | 8.46 | 5.14 | 0 | 9.16 | 22.76 | 27.59 | 13.29 |
| **Standard care vs Stress Echo** | 14.49 | 1.89 | 12.6 | 0 | 13.91 | 28.4 | 1.89 | 26.82 |

Table 2: Pairwise Comparisons Table for the network of 18 antidepressants

|  | **Number of studies in each comparison** | | **“known unknowns”** | **“unknown unknowns”** | **Overall bias** |
| --- | --- | --- | --- | --- | --- |
| **Pairwise comparison** | **Reporting this outcome (sample size)** | **Total identified in the SR (total sample size)** | **Classification system & signalling questions** | **Qualitative signals & quantitative considerations** | **Synthesizing judgements** |
| **Group A:  observed for this outcome** | | | | | |
| **Agomelatine Vs. Duloxetine** | 1 ( 418 ) | 1 ( 418 ) | Undetected bias | Undetected bias | Undetected bias |
| **Agomelatine Vs. Escitalopram** | 2 ( 462 ) | 2 ( 462 ) | Undetected bias | Undetected bias | Undetected bias |
| **Agomelatine Vs. Fluoxetine** | 2 ( 1143 ) | 2 ( 1143 ) | Undetected bias | Undetected bias | Undetected bias |
| **Agomelatine Vs. Paroxetine** | 2 ( 747 ) | 2 ( 747 ) | Undetected bias | Undetected bias | Undetected bias |
| **Agomelatine Vs. Venlafaxine** | 2 ( 609 ) | 2 ( 609 ) | Undetected bias | Undetected bias | Undetected bias |
| **Amitriptyline Vs. Fluoxetine** | 12 ( 888 ) | 14 ( 979 ) | Suspected bias favouring Fluoxetine | Suspected bias favouring Fluoxetine | Suspected bias favouring Fluoxetine |
| **Amitriptyline Vs. Fluvoxamine** | 3 ( 337 ) | 3 ( 337 ) | Undetected bias | Suspected bias favouring Fluvoxamine | Suspected bias favouring Fluvoxamine |
| **Amitriptyline Vs. Milnacipran** | 2 ( 231 ) | 2 ( 231 ) | Undetected bias | Suspected bias favouring Milnacipran | Suspected bias favouring Milnacipran |
| **Amitriptyline Vs. Paroxetine** | 13 ( 1575 ) | 18 ( 1688 ) | Undetected bias | Undetected bias | Undetected bias |
| **Amitriptyline Vs. Sertraline** | 4 ( 709 ) | 4 ( 709 ) | Undetected bias | Suspected bias favouring Sertraline | Suspected bias favouring Sertraline |
| **Amitriptyline Vs. Trazodone** | 2 ( 229 ) | 3 ( 269 ) | Suspected bias favouring Trazodone | Suspected bias favouring Trazodone | Suspected bias favouring Trazodone |
| **Amitriptyline Vs. Venlafaxine** | 2 ( 272 ) | 2 ( 272 ) | Undetected bias | Suspected bias favouring Venlafaxine | Suspected bias favouring Venlafaxine |
| **Bupropion Vs. Fluoxetine** | 1 ( 123 ) | 1 ( 123 ) | Undetected bias | Undetected bias | Undetected bias |
| **Bupropion Vs. Paroxetine** | 1 ( 140 ) | 2 ( 218 ) | Undetected bias | Undetected bias | Undetected bias |
| **Bupropion Vs. Sertraline** | 1 ( 16 ) | 1 ( 16 ) | Undetected bias | Suspected bias favouring Sertraline | Suspected bias favouring Sertraline |
| **Bupropion Vs. Trazodone** | 1 ( 124 ) | 1 ( 124 ) | Undetected bias | Suspected bias favouring Trazodone | Suspected bias favouring Trazodone |
| **Bupropion Vs. Venlafaxine** | 1 ( 348 ) | 1 ( 348 ) | Undetected bias | Suspected bias favouring Venlafaxine | Suspected bias favouring Venlafaxine |
| **Citalopram Vs. Clomipramine** | 1 ( 114 ) | 1 ( 114 ) | Undetected bias | Suspected bias favouring Citalopram | Suspected bias favouring Citalopram |
| **Citalopram Vs. Escitalopram** | 10 ( 1511 ) | 10 ( 1511 ) | Undetected bias | Suspected bias favouring Escitalopram | Suspected bias favouring Escitalopram |
| **Citalopram Vs. Fluoxetine** | 2 ( 673 ) | 2 ( 673 ) | Undetected bias | Suspected bias favouring Citalopram | Suspected bias favouring Citalopram |
| **Citalopram Vs. Fluvoxamine** | 1 ( 217 ) | 1 ( 217 ) | Undetected bias | Suspected bias favouring Citalopram | Suspected bias favouring Citalopram |
| **Citalopram Vs. Mirtazapine** | 1 ( 270 ) | 2 ( 322 ) | Suspected bias favouring Mirtazapine | Suspected bias favouring Mirtazapine | Suspected bias favouring Mirtazapine |
| **Citalopram Vs. Reboxetine** | 1 ( 359 ) | 1 ( 359 ) | Undetected bias | Suspected bias favouring Reboxetine | Suspected bias favouring Reboxetine |
| **Citalopram Vs. Sertraline** | 3 ( 596 ) | 3 ( 596 ) | Undetected bias | Suspected bias favouring Citalopram | Suspected bias favouring Citalopram |
| **Citalopram Vs. Venlafaxine** | 2 ( 204 ) | 2 ( 204 ) | Undetected bias | Suspected bias favouring Venlafaxine | Suspected bias favouring Venlafaxine |
| **Clomipramine Vs. Fluoxetine** | 4 ( 347 ) | 4 ( 347 ) | Undetected bias | Suspected bias favouring Fluoxetine | Suspected bias favouring Fluoxetine |
| **Clomipramine Vs. Fluvoxamine** | 2 ( 83 ) | 2 ( 83 ) | Undetected bias | Suspected bias favouring Fluvoxamine | Suspected bias favouring Fluvoxamine |
| **Clomipramine Vs. Milnacipran** | 1 ( 107 ) | 1 ( 107 ) | Undetected bias | Suspected bias favouring Milnacipran | Suspected bias favouring Milnacipran |
| **Clomipramine Vs. Paroxetine** | 6 ( 1455 ) | 6 ( 1455 ) | Undetected bias | Undetected bias | Undetected bias |
| **Clomipramine Vs. Sertraline** | 2 ( 272 ) | 2 ( 272 ) | Undetected bias | Suspected bias favouring Sertraline | Suspected bias favouring Sertraline |
| **Clomipramine Vs. Trazodone** | 1 ( 115 ) | 1 ( 115 ) | Undetected bias | Suspected bias favouring Trazodone | Suspected bias favouring Trazodone |
| **Clomipramine Vs. Venlafaxine** | 2 ( 215 ) | 2 ( 215 ) | Undetected bias | Suspected bias favouring Venlafaxine | Suspected bias favouring Venlafaxine |
| **Duloxetine Vs. Escitalopram** | 2 ( 573 ) | 2 ( 573 ) | Undetected bias | Suspected bias favouring Duloxetine | Suspected bias favouring Duloxetine |
| **Duloxetine Vs. Paroxetine** | 2 ( 759 ) | 2 ( 759 ) | Undetected bias | Suspected bias favouring Duloxetine | Suspected bias favouring Duloxetine |
| **Duloxetine Vs. Venlafaxine** | 2 ( 836 ) | 2 ( 836 ) | Undetected bias | Suspected bias favouring Duloxetine | Suspected bias favouring Duloxetine |
| **Escitalopram Vs. Fluoxetine** | 2 ( 445 ) | 3 ( 475 ) | Suspected bias favouring Escitalopram | Suspected bias favouring Escitalopram | Suspected bias favouring Escitalopram |
| **Escitalopram Vs. Paroxetine** | 2 ( 784 ) | 2 ( 784 ) | Undetected bias | Suspected bias favouring Escitalopram | Suspected bias favouring Escitalopram |
| **Escitalopram Vs. Sertraline** | 2 ( 355 ) | 2 ( 355 ) | Undetected bias | Suspected bias favouring Escitalopram | Suspected bias favouring Escitalopram |
| **Escitalopram Vs. Venlafaxine** | 2 ( 495 ) | 2 ( 495 ) | Undetected bias | Suspected bias favouring Escitalopram | Suspected bias favouring Escitalopram |
| **Fluoxetine Vs. Fluvoxamine** | 2 ( 284 ) | 2 ( 284 ) | Undetected bias | Suspected bias favouring Fluoxetine | Suspected bias favouring Fluoxetine |
| **Fluoxetine Vs. Milnacipran** | 2 ( 490 ) | 2 ( 490 ) | Undetected bias | Suspected bias favouring Milnacipran | Suspected bias favouring Milnacipran |
| **Fluoxetine Vs. Mirtazapine** | 5 ( 615 ) | 5 ( 615 ) | Undetected bias | Suspected bias favouring Mirtazapine | Suspected bias favouring Mirtazapine |
| **Fluoxetine Vs. Nefazodone** | 3 ( 161 ) | 3 ( 161 ) | Undetected bias | Suspected bias favouring Nefazodone | Suspected bias favouring Nefazodone |
| **Fluoxetine Vs. Paroxetine** | 9 ( 1364 ) | 10 ( 1385 ) | Undetected bias | Undetected bias | Undetected bias |
| **Fluoxetine Vs. Reboxetine** | 2 ( 253 ) | 2 ( 253 ) | Undetected bias | Suspected bias favouring Reboxetine | Suspected bias favouring Reboxetine |
| **Fluoxetine Vs. Sertraline** | 6 ( 1221 ) | 6 ( 1221 ) | Undetected bias | Suspected bias favouring Sertraline | Suspected bias favouring Sertraline |
| **Fluoxetine Vs. Trazodone** | 4 ( 234 ) | 4 ( 234 ) | Undetected bias | Suspected bias favouring Trazodone | Suspected bias favouring Trazodone |
| **Fluoxetine Vs. Venlafaxine** | 9 ( 2538 ) | 10 ( 2804 ) | Suspected bias favouring Venlafaxine | Suspected bias favouring Venlafaxine | Suspected bias favouring Venlafaxine |
| **Fluvoxamine Vs. Milnacipran** | 2 ( 239 ) | 2 ( 239 ) | Undetected bias | Suspected bias favouring Milnacipran | Suspected bias favouring Milnacipran |
| **Fluvoxamine Vs. Mirtazapine** | 2 ( 412 ) | 2 ( 412 ) | Undetected bias | Suspected bias favouring Mirtazapine | Suspected bias favouring Mirtazapine |
| **Fluvoxamine Vs. Paroxetine** | 2 ( 180 ) | 2 ( 180 ) | Undetected bias | Undetected bias | Undetected bias |
| **Fluvoxamine Vs. Sertraline** | 2 ( 185 ) | 2 ( 185 ) | Undetected bias | Suspected bias favouring Sertraline | Suspected bias favouring Sertraline |
| **Fluvoxamine Vs. Venlafaxine** | 1 ( 111 ) | 1 ( 111 ) | Undetected bias | Suspected bias favouring Venlafaxine | Suspected bias favouring Venlafaxine |
| **Milnacipran Vs. Paroxetine** | 2 ( 1207 ) | 2 ( 1207 ) | Undetected bias | Suspected bias favouring Milnacipran | Suspected bias favouring Milnacipran |
| **Milnacipran Vs. Sertraline** | 1 ( 53 ) | 1 ( 53 ) | Undetected bias | Suspected bias favouring Milnacipran | Suspected bias favouring Milnacipran |
| **Mirtazapine Vs. Paroxetine** | 5 ( 916 ) | 5 ( 916 ) | Undetected bias | Suspected bias favouring Mirtazapine | Suspected bias favouring Mirtazapine |
| **Mirtazapine Vs. Sertraline** | 1 ( 346 ) | 1 ( 346 ) | Undetected bias | Suspected bias favouring Mirtazapine | Suspected bias favouring Mirtazapine |
| **Mirtazapine Vs. Trazodone** | 1 ( 200 ) | 1 ( 200 ) | Undetected bias | Suspected bias favouring Mirtazapine | Suspected bias favouring Mirtazapine |
| **Mirtazapine Vs. Venlafaxine** | 2 ( 533 ) | 2 ( 533 ) | Undetected bias | Suspected bias favouring Venlafaxine | Suspected bias favouring Venlafaxine |
| **Nefazodone Vs. Paroxetine** | 2 ( 246 ) | 2 ( 246 ) | Undetected bias | Suspected bias favouring Nefazodone | Suspected bias favouring Nefazodone |
| **Nefazodone Vs. Sertraline** | 1 ( 160 ) | 1 ( 160 ) | Undetected bias | Suspected bias favouring Nefazodone | Suspected bias favouring Nefazodone |
| **Paroxetine Vs. Reboxetine** | 1 ( 325 ) | 1 ( 325 ) | Undetected bias | Suspected bias favouring Reboxetine | Suspected bias favouring Reboxetine |
| **Paroxetine Vs. Sertraline** | 2 ( 545 ) | 2 ( 545 ) | Undetected bias | Undetected bias | Undetected bias |
| **Paroxetine Vs. Trazodone** | 2 ( 333 ) | 2 ( 333 ) | Undetected bias | Undetected bias | Undetected bias |
| **Paroxetine Vs. Venlafaxine** | 2 ( 475 ) | 2 ( 475 ) | Undetected bias | Suspected bias favouring Venlafaxine | Suspected bias favouring Venlafaxine |
| **Reboxetine Vs. Venlafaxine** | 1 ( 167 ) | 1 ( 167 ) | Undetected bias | Suspected bias favouring Venlafaxine | Suspected bias favouring Venlafaxine |
| **Sertraline Vs. Trazodone** | 1 ( 122 ) | 1 ( 122 ) | Undetected bias | Suspected bias favouring Sertraline | Suspected bias favouring Sertraline |
| **Sertraline Vs. Venlafaxine** | 3 ( 470 ) | 3 ( 470 ) | Undetected bias | Suspected bias favouring Venlafaxine | Suspected bias favouring Venlafaxine |
| **Trazodone Vs. Venlafaxine** | 1 ( 112 ) | 1 ( 112 ) | Undetected bias | Suspected bias favouring Venlafaxine | Suspected bias favouring Venlafaxine |
| **Venlafaxine Vs. Vortioxetine** | 1 ( 443 ) | 1 ( 443 ) | Undetected bias | Undetected bias | Undetected bias |
| **Group B: observed for other outcomes** | | | | | |
| **Amitriptyline Vs. Bupropion** | 0 ( 0 ) | 1 ( 118 ) | Undetected bias | Undetected bias | Undetected bias |
| **Amitriptyline Vs. Nefazodone** | 0 ( 0 ) | 1 ( 37 ) | Undetected bias | Suspected bias favouring Nefazodone | Suspected bias favouring Nefazodone |
| **Group C: Unobserved** | | | | | |
| **Agomelatine Vs. Amitriptyline** | 0 ( 0 ) | 0 ( 0 ) | NA | Undetected bias | Undetected bias |
| **Agomelatine Vs. Bupropion** | 0 ( 0 ) | 0 ( 0 ) | NA | Undetected bias | Undetected bias |
| **Agomelatine Vs. Citalopram** | 0 ( 0 ) | 0 ( 0 ) | NA | Undetected bias | Undetected bias |
| **Agomelatine Vs. Clomipramine** | 0 ( 0 ) | 0 ( 0 ) | NA | Undetected bias | Undetected bias |
| **Agomelatine Vs. Fluvoxamine** | 0 ( 0 ) | 0 ( 0 ) | NA | Undetected bias | Undetected bias |
| **Agomelatine Vs. Milnacipran** | 0 ( 0 ) | 0 ( 0 ) | NA | Undetected bias | Undetected bias |
| **Agomelatine Vs. Mirtazapine** | 0 ( 0 ) | 0 ( 0 ) | NA | Undetected bias | Undetected bias |
| **Agomelatine Vs. Nefazodone** | 0 ( 0 ) | 0 ( 0 ) | NA | Undetected bias | Undetected bias |
| **Agomelatine Vs. Reboxetine** | 0 ( 0 ) | 0 ( 0 ) | NA | Undetected bias | Undetected bias |
| **Agomelatine Vs. Sertraline** | 0 ( 0 ) | 0 ( 0 ) | NA | Undetected bias | Undetected bias |
| **Agomelatine Vs. Trazodone** | 0 ( 0 ) | 0 ( 0 ) | NA | Undetected bias | Undetected bias |
| **Agomelatine Vs. Vortioxetine** | 0 ( 0 ) | 0 ( 0 ) | NA | Undetected bias | Undetected bias |
| **Amitriptyline Vs. Citalopram** | 0 ( 0 ) | 0 ( 0 ) | NA | Suspected bias favouring Citalopram | Suspected bias favouring Citalopram |
| **Amitriptyline Vs. Clomipramine** | 0 ( 0 ) | 0 ( 0 ) | NA | Undetected bias | Undetected bias |
| **Amitriptyline Vs. Duloxetine** | 0 ( 0 ) | 0 ( 0 ) | NA | Suspected bias favouring Duloxetine | Suspected bias favouring Duloxetine |
| **Amitriptyline Vs. Escitalopram** | 0 ( 0 ) | 0 ( 0 ) | NA | Suspected bias favouring Escitalopram | Suspected bias favouring Escitalopram |
| **Amitriptyline Vs. Mirtazapine** | 0 ( 0 ) | 0 ( 0 ) | NA | Suspected bias favouring Mirtazapine | Suspected bias favouring Mirtazapine |
| **Amitriptyline Vs. Reboxetine** | 0 ( 0 ) | 0 ( 0 ) | NA | Suspected bias favouring Reboxetine | Suspected bias favouring Reboxetine |
| **Amitriptyline Vs. Vortioxetine** | 0 ( 0 ) | 0 ( 0 ) | NA | Undetected bias | Undetected bias |
| **Bupropion Vs. Citalopram** | 0 ( 0 ) | 0 ( 0 ) | NA | Suspected bias favouring Citalopram | Suspected bias favouring Citalopram |
| **Bupropion Vs. Clomipramine** | 0 ( 0 ) | 0 ( 0 ) | NA | Undetected bias | Undetected bias |
| **Bupropion Vs. Duloxetine** | 0 ( 0 ) | 0 ( 0 ) | NA | Suspected bias favouring Duloxetine | Suspected bias favouring Duloxetine |
| **Bupropion Vs. Escitalopram** | 0 ( 0 ) | 0 ( 0 ) | NA | Suspected bias favouring Escitalopram | Suspected bias favouring Escitalopram |
| **Bupropion Vs. Fluvoxamine** | 0 ( 0 ) | 0 ( 0 ) | NA | Suspected bias favouring Fluvoxamine | Suspected bias favouring Fluvoxamine |
| **Bupropion Vs. Milnacipran** | 0 ( 0 ) | 0 ( 0 ) | NA | Suspected bias favouring Milnacipran | Suspected bias favouring Milnacipran |
| **Bupropion Vs. Mirtazapine** | 0 ( 0 ) | 0 ( 0 ) | NA | Suspected bias favouring Mirtazapine | Suspected bias favouring Mirtazapine |
| **Bupropion Vs. Nefazodone** | 0 ( 0 ) | 0 ( 0 ) | NA | Suspected bias favouring Nefazodone | Suspected bias favouring Nefazodone |
| **Bupropion Vs. Reboxetine** | 0 ( 0 ) | 0 ( 0 ) | NA | Suspected bias favouring Reboxetine | Suspected bias favouring Reboxetine |
| **Bupropion Vs. Vortioxetine** | 0 ( 0 ) | 0 ( 0 ) | NA | Undetected bias | Undetected bias |
| **Citalopram Vs. Duloxetine** | 0 ( 0 ) | 0 ( 0 ) | NA | Suspected bias favouring Duloxetine | Suspected bias favouring Duloxetine |
| **Citalopram Vs. Milnacipran** | 0 ( 0 ) | 0 ( 0 ) | NA | Suspected bias favouring Milnacipran | Suspected bias favouring Milnacipran |
| **Citalopram Vs. Nefazodone** | 0 ( 0 ) | 0 ( 0 ) | NA | Suspected bias favouring Nefazodone | Suspected bias favouring Nefazodone |
| **Citalopram Vs. Paroxetine** | 0 ( 0 ) | 0 ( 0 ) | NA | Undetected bias | Undetected bias |
| **Citalopram Vs. Trazodone** | 0 ( 0 ) | 0 ( 0 ) | NA | Suspected bias favouring Trazodone | Suspected bias favouring Trazodone |
| **Citalopram Vs. Vortioxetine** | 0 ( 0 ) | 0 ( 0 ) | NA | Undetected bias | Undetected bias |
| **Clomipramine Vs. Duloxetine** | 0 ( 0 ) | 0 ( 0 ) | NA | Suspected bias favouring Duloxetine | Suspected bias favouring Duloxetine |
| **Clomipramine Vs. Escitalopram** | 0 ( 0 ) | 0 ( 0 ) | NA | Suspected bias favouring Escitalopram | Suspected bias favouring Escitalopram |
| **Clomipramine Vs. Mirtazapine** | 0 ( 0 ) | 0 ( 0 ) | NA | Suspected bias favouring Mirtazapine | Suspected bias favouring Mirtazapine |
| **Clomipramine Vs. Nefazodone** | 0 ( 0 ) | 0 ( 0 ) | NA | Suspected bias favouring Nefazodone | Suspected bias favouring Nefazodone |
| **Clomipramine Vs. Reboxetine** | 0 ( 0 ) | 0 ( 0 ) | NA | Suspected bias favouring Reboxetine | Suspected bias favouring Reboxetine |
| **Clomipramine Vs. Vortioxetine** | 0 ( 0 ) | 0 ( 0 ) | NA | Undetected bias | Undetected bias |
| **Duloxetine Vs. Fluoxetine** | 0 ( 0 ) | 0 ( 0 ) | NA | Suspected bias favouring Duloxetine | Suspected bias favouring Duloxetine |
| **Duloxetine Vs. Fluvoxamine** | 0 ( 0 ) | 0 ( 0 ) | NA | Suspected bias favouring Duloxetine | Suspected bias favouring Duloxetine |
| **Duloxetine Vs. Milnacipran** | 0 ( 0 ) | 0 ( 0 ) | NA | Suspected bias favouring Duloxetine | Suspected bias favouring Duloxetine |
| **Duloxetine Vs. Mirtazapine** | 0 ( 0 ) | 0 ( 0 ) | NA | Suspected bias favouring Duloxetine | Suspected bias favouring Duloxetine |
| **Duloxetine Vs. Nefazodone** | 0 ( 0 ) | 0 ( 0 ) | NA | Suspected bias favouring Duloxetine | Suspected bias favouring Duloxetine |
| **Duloxetine Vs. Reboxetine** | 0 ( 0 ) | 0 ( 0 ) | NA | Suspected bias favouring Duloxetine | Suspected bias favouring Duloxetine |
| **Duloxetine Vs. Sertraline** | 0 ( 0 ) | 0 ( 0 ) | NA | Suspected bias favouring Duloxetine | Suspected bias favouring Duloxetine |
| **Duloxetine Vs. Trazodone** | 0 ( 0 ) | 0 ( 0 ) | NA | Suspected bias favouring Duloxetine | Suspected bias favouring Duloxetine |
| **Duloxetine Vs. Vortioxetine** | 0 ( 0 ) | 0 ( 0 ) | NA | Undetected bias | Undetected bias |
| **Escitalopram Vs. Fluvoxamine** | 0 ( 0 ) | 0 ( 0 ) | NA | Suspected bias favouring Escitalopram | Suspected bias favouring Escitalopram |
| **Escitalopram Vs. Milnacipran** | 0 ( 0 ) | 0 ( 0 ) | NA | Suspected bias favouring Escitalopram | Suspected bias favouring Escitalopram |
| **Escitalopram Vs. Mirtazapine** | 0 ( 0 ) | 0 ( 0 ) | NA | Suspected bias favouring Escitalopram | Suspected bias favouring Escitalopram |
| **Escitalopram Vs. Nefazodone** | 0 ( 0 ) | 0 ( 0 ) | NA | Suspected bias favouring Escitalopram | Suspected bias favouring Escitalopram |
| **Escitalopram Vs. Reboxetine** | 0 ( 0 ) | 0 ( 0 ) | NA | Suspected bias favouring Escitalopram | Suspected bias favouring Escitalopram |
| **Escitalopram Vs. Trazodone** | 0 ( 0 ) | 0 ( 0 ) | NA | Suspected bias favouring Escitalopram | Suspected bias favouring Escitalopram |
| **Escitalopram Vs. Vortioxetine** | 0 ( 0 ) | 0 ( 0 ) | NA | Undetected bias | Undetected bias |
| **Fluoxetine Vs. Vortioxetine** | 0 ( 0 ) | 0 ( 0 ) | NA | Undetected bias | Undetected bias |
| **Fluvoxamine Vs. Nefazodone** | 0 ( 0 ) | 0 ( 0 ) | NA | Suspected bias favouring Nefazodone | Suspected bias favouring Nefazodone |
| **Fluvoxamine Vs. Reboxetine** | 0 ( 0 ) | 0 ( 0 ) | NA | Suspected bias favouring Reboxetine | Suspected bias favouring Reboxetine |
| **Fluvoxamine Vs. Trazodone** | 0 ( 0 ) | 0 ( 0 ) | NA | Suspected bias favouring Fluvoxamine | Suspected bias favouring Fluvoxamine |
| **Fluvoxamine Vs. Vortioxetine** | 0 ( 0 ) | 0 ( 0 ) | NA | Undetected bias | Undetected bias |
| **Milnacipran Vs. Mirtazapine** | 0 ( 0 ) | 0 ( 0 ) | NA | Suspected bias favouring Milnacipran | Suspected bias favouring Milnacipran |
| **Milnacipran Vs. Nefazodone** | 0 ( 0 ) | 0 ( 0 ) | NA | Suspected bias favouring Milnacipran | Suspected bias favouring Milnacipran |
| **Milnacipran Vs. Reboxetine** | 0 ( 0 ) | 0 ( 0 ) | NA | Suspected bias favouring Reboxetine | Suspected bias favouring Reboxetine |
| **Milnacipran Vs. Trazodone** | 0 ( 0 ) | 0 ( 0 ) | NA | Suspected bias favouring Trazodone | Suspected bias favouring Trazodone |
| **Milnacipran Vs. Venlafaxine** | 0 ( 0 ) | 0 ( 0 ) | NA | Suspected bias favouring Venlafaxine | Suspected bias favouring Venlafaxine |
| **Milnacipran Vs. Vortioxetine** | 0 ( 0 ) | 0 ( 0 ) | NA | Undetected bias | Undetected bias |
| **Mirtazapine Vs. Nefazodone** | 0 ( 0 ) | 0 ( 0 ) | NA | Suspected bias favouring Nefazodone | Suspected bias favouring Nefazodone |
| **Mirtazapine Vs. Reboxetine** | 0 ( 0 ) | 0 ( 0 ) | NA | Suspected bias favouring Reboxetine | Suspected bias favouring Reboxetine |
| **Mirtazapine Vs. Vortioxetine** | 0 ( 0 ) | 0 ( 0 ) | NA | Undetected bias | Undetected bias |
| **Nefazodone Vs. Reboxetine** | 0 ( 0 ) | 0 ( 0 ) | NA | Suspected bias favouring Reboxetine | Suspected bias favouring Reboxetine |
| **Nefazodone Vs. Trazodone** | 0 ( 0 ) | 0 ( 0 ) | NA | Suspected bias favouring Nefazodone | Suspected bias favouring Nefazodone |
| **Nefazodone Vs. Venlafaxine** | 0 ( 0 ) | 0 ( 0 ) | NA | Suspected bias favouring Venlafaxine | Suspected bias favouring Venlafaxine |
| **Nefazodone Vs. Vortioxetine** | 0 ( 0 ) | 0 ( 0 ) | NA | Undetected bias | Undetected bias |
| **Paroxetine Vs. Vortioxetine** | 0 ( 0 ) | 0 ( 0 ) | NA | Undetected bias | Undetected bias |
| **Reboxetine Vs. Sertraline** | 0 ( 0 ) | 0 ( 0 ) | NA | Suspected bias favouring Reboxetine | Suspected bias favouring Reboxetine |
| **Reboxetine Vs. Trazodone** | 0 ( 0 ) | 0 ( 0 ) | NA | Suspected bias favouring Reboxetine | Suspected bias favouring Reboxetine |
| **Reboxetine Vs. Vortioxetine** | 0 ( 0 ) | 0 ( 0 ) | NA | Undetected bias | Undetected bias |
| **Sertraline Vs. Vortioxetine** | 0 ( 0 ) | 0 ( 0 ) | NA | Undetected bias | Undetected bias |
| **Trazodone Vs. Vortioxetine** | 0 ( 0 ) | 0 ( 0 ) | NA | Undetected bias | Undetected bias |

Table 3: ROB-MEN Table for the network of 18 antidepressants

|  | **1** | **2** | **3** | **4** | **5** | **6** | **7** | | **8** | | | **9** | | |
| --- | --- | --- | --- | --- | --- | --- | --- | --- | --- | --- | --- | --- | --- | --- |
|  | **% contribution of evidence from pairwise comparisons with suspected bias** | | | **Evaluation of contribution from evidence with suspected bias** | **Bias assessment for indirect evidence** | **NMA treatment effect** | **NMR treatment effect at the smallest observed variance** | | **Evaluation of small-study effects** | | | **Qualitative merging / overall bias** | | |
| **NMA estimate** | **Favouring first treatment** | **Favouring second treatment** | **Total from all comparisons** |  |  |  |  |  |  |  |  |  |  |  |
| **Mixed/ only direct** | | | | | | | | | | | | | | |
| **Agomelatine Vs. Duloxetine** | 0.00 | 32.38 | 38.24 | Substantial contribution from bias favouring Duloxetine |  | 1.05 (0.81, 1.37) | | 1.03 (0.75, 1.37) | | No evidence of small-study effects | | | Some concerns | |
| **Agomelatine Vs. Escitalopram** | 0.00 | 25.36 | 41.77 | Substantial contribution from bias favouring Escitalopram |  | 0.9 (0.71, 1.14) | | 0.89 (0.67, 1.17) | | No evidence of small-study effects | | | Some concerns | |
| **Agomelatine Vs. Fluoxetine** | 0.00 | 3.37 | 32.58 | No substantial contribution from bias |  | 1.2 (0.98, 1.47) | | 1.17 (0.94, 1.46) | | No evidence of small-study effects | | | Low risk | |
| **Agomelatine Vs. Paroxetine** | 0.00 | 0.00 | 29.26 | No substantial contribution from bias |  | 1.01 (0.82, 1.24) | | 1.01 (0.79, 1.27) | | No evidence of small-study effects | | | Low risk | |
| **Agomelatine Vs. Venlafaxine** | 0.00 | 22.37 | 35.92 | Substantial contribution from bias favouring Venlafaxine |  | 1.01 (0.81, 1.25) | | 1 (0.78, 1.27) | | No evidence of small-study effects | | | Some concerns | |
| **Amitriptyline Vs. Fluoxetine** | 0.00 | 39.66 | 78.19 | Substantial contribution from bias favouring Fluoxetine |  | 1.25 (1.06, 1.47) | | 1.29 (1.05, 1.58) | | No evidence of small-study effects | | | Some concerns | |
| **Amitriptyline Vs. Fluvoxamine** | 0.00 | 20.51 | 83.40 | Substantial contribution from bias favouring Fluvoxamine |  | 1.26 (0.99, 1.6) | | 1.31 (0.96, 1.79) | | No evidence of small-study effects | | | Some concerns | |
| **Amitriptyline Vs. Milnacipran** | 0.00 | 55.18 | 82.35 | Substantial contribution from bias favouring Milnacipran |  | 1.12 (0.87, 1.44) | | 1.17 (0.88, 1.55) | | No evidence of small-study effects | | | Some concerns | |
| **Amitriptyline Vs. Paroxetine** | 0.00 | 0.00 | 40.57 | No substantial contribution from bias |  | 1.05 (0.9, 1.23) | | 1.11 (0.92, 1.33) | | No evidence of small-study effects | | | Low risk | |
| **Amitriptyline Vs. Sertraline** | 0.00 | 37.56 | 81.17 | Substantial contribution from bias favouring Sertraline |  | 1.12 (0.93, 1.35) | | 1.12 (0.9, 1.4) | | No evidence of small-study effects | | | Some concerns | |
| **Amitriptyline Vs. Trazodone** | 0.00 | 30.10 | 70.63 | Substantial contribution from bias favouring Trazodone |  | 1.41 (1.07, 1.85) | | 1.42 (1.02, 1.95) | | No evidence of small-study effects | | | Some concerns | |
| **Amitriptyline Vs. Venlafaxine** | 0.00 | 43.97 | 82.12 | Substantial contribution from bias favouring Venlafaxine |  | 1.06 (0.87, 1.27) | | 1.1 (0.87, 1.38) | | No evidence of small-study effects | | | Some concerns | |
| **Bupropion Vs. Fluoxetine** | 0.00 | 4.10 | 53.54 | No substantial contribution from bias |  | 1.37 (0.96, 1.96) | | 1.39 (0.95, 2.05) | | No evidence of small-study effects | | | Low risk | |
| **Bupropion Vs. Paroxetine** | 0.00 | 0.00 | 44.18 | No substantial contribution from bias |  | 1.15 (0.8, 1.67) | | 1.19 (0.81, 1.76) | | No evidence of small-study effects | | | Low risk | |
| **Bupropion Vs. Sertraline** | 0.00 | 22.15 | 69.72 | Substantial contribution from bias favouring Sertraline |  | 1.23 (0.84, 1.8) | | 1.2 (0.81, 1.81) | | No evidence of small-study effects | | | Some concerns | |
| **Bupropion Vs. Trazodone** | 0.00 | 30.58 | 65.67 | Substantial contribution from bias favouring Trazodone |  | 1.54 (1.03, 2.31) | | 1.53 (0.97, 2.4) | | No evidence of small-study effects | | | Some concerns | |
| **Bupropion Vs. Venlafaxine** | 0.00 | 59.25 | 74.38 | Substantial contribution from bias favouring Venlafaxine |  | 1.16 (0.81, 1.66) | | 1.18 (0.81, 1.74) | | No evidence of small-study effects | | | Some concerns | |
| **Citalopram Vs. Clomipramine** | 23.78 | 0.00 | 85.64 | Substantial contribution from bias favouring Citalopram |  | 1.06 (0.83, 1.38) | | 0.98 (0.7, 1.34) | | No evidence of small-study effects | | | Some concerns | |
| **Citalopram Vs. Escitalopram** | 9.53 | 77.74 | 95.89 | Substantial contribution from bias favouring Escitalopram |  | 0.8 (0.65, 0.97) | | 0.77 (0.61, 0.98) | | No evidence of small-study effects | | | Some concerns | |
| **Citalopram Vs. Fluoxetine** | 30.23 | 4.53 | 91.49 | Substantial contribution from bias favouring Citalopram |  | 1.06 (0.87, 1.29) | | 1.02 (0.82, 1.26) | | No evidence of small-study effects | | | Some concerns | |
| **Citalopram Vs. Fluvoxamine** | 24.23 | 6.68 | 93.72 | Substantial contribution from bias favouring Citalopram |  | 1.07 (0.82, 1.39) | | 1.03 (0.74, 1.45) | | No evidence of small-study effects | | | Some concerns | |
| **Citalopram Vs. Mirtazapine** | 16.42 | 45.62 | 95.79 | Substantial contribution from bias favouring Mirtazapine |  | 0.83 (0.65, 1.06) | | 0.78 (0.59, 1.01) | | No evidence of small-study effects | | | Some concerns | |
| **Citalopram Vs. Reboxetine** | 12.37 | 46.28 | 95.51 | Substantial contribution from bias favouring Reboxetine |  | 1.28 (0.92, 1.75) | | 1.23 (0.86, 1.78) | | No evidence of small-study effects | | | Some concerns | |
| **Citalopram Vs. Sertraline** | 36.53 | 13.34 | 95.17 | Substantial contribution from bias favouring Citalopram |  | 0.95 (0.77, 1.19) | | 0.88 (0.7, 1.12) | | No evidence of small-study effects | | | Some concerns | |
| **Citalopram Vs. Venlafaxine** | 17.07 | 37.98 | 94.02 | Substantial contribution from bias favouring Venlafaxine |  | 0.9 (0.73, 1.11) | | 0.87 (0.67, 1.11) | | No evidence of small-study effects | | | Some concerns | |
| **Clomipramine Vs. Fluoxetine** | 0.00 | 27.60 | 75.22 | Substantial contribution from bias favouring Fluoxetine |  | 1 (0.82, 1.22) | | 1.04 (0.8, 1.38) | | No evidence of small-study effects | | | Some concerns | |
| **Clomipramine Vs. Fluvoxamine** | 0.00 | 22.19 | 82.33 | Substantial contribution from bias favouring Fluvoxamine |  | 1.01 (0.76, 1.32) | | 1.06 (0.74, 1.53) | | No evidence of small-study effects | | | Some concerns | |
| **Clomipramine Vs. Milnacipran** | 0.00 | 51.29 | 81.54 | Substantial contribution from bias favouring Milnacipran |  | 0.9 (0.67, 1.19) | | 0.95 (0.67, 1.33) | | No evidence of small-study effects | | | Some concerns | |
| **Clomipramine Vs. Paroxetine** | 0.00 | 0.00 | 42.79 | No substantial contribution from bias |  | 0.84 (0.68, 1.02) | | 0.9 (0.69, 1.17) | | No evidence of small-study effects | | | Low risk | |
| **Clomipramine Vs. Sertraline** | 0.00 | 34.04 | 79.52 | Substantial contribution from bias favouring Sertraline |  | 0.89 (0.71, 1.12) | | 0.9 (0.68, 1.22) | | No evidence of small-study effects | | | Some concerns | |
| **Clomipramine Vs. Trazodone** | 0.00 | 26.57 | 69.59 | Substantial contribution from bias favouring Trazodone |  | 1.13 (0.81, 1.52) | | 1.15 (0.79, 1.65) | | No evidence of small-study effects | | | Some concerns | |
| **Clomipramine Vs. Venlafaxine** | 0.00 | 44.85 | 80.65 | Substantial contribution from bias favouring Venlafaxine |  | 0.84 (0.67, 1.06) | | 0.89 (0.66, 1.2) | | No evidence of small-study effects | | | Some concerns | |
| **Duloxetine Vs. Escitalopram** | 49.74 | 21.08 | 82.01 | Substantial contribution from bias favouring Duloxetine |  | 0.85 (0.67, 1.08) | | 0.86 (0.65, 1.18) | | No evidence of small-study effects | | | Some concerns | |
| **Duloxetine Vs. Paroxetine** | 53.40 | 0.00 | 77.31 | Substantial contribution from bias favouring Duloxetine |  | 0.96 (0.76, 1.2) | | 0.98 (0.76, 1.29) | | No evidence of small-study effects | | | Some concerns | |
| **Duloxetine Vs. Venlafaxine** | 49.14 | 17.37 | 78.82 | Substantial contribution from bias favouring Duloxetine |  | 0.96 (0.77, 1.21) | | 0.97 (0.74, 1.29) | | No evidence of small-study effects | | | Some concerns | |
| **Escitalopram Vs. Fluoxetine** | 45.08 | 3.66 | 82.92 | Substantial contribution from bias favouring Escitalopram |  | 1.33 (1.11, 1.6) | | 1.32 (1.06, 1.63) | | No evidence of small-study effects | | | Some concerns | |
| **Escitalopram Vs. Paroxetine** | 42.83 | 0.00 | 73.64 | Substantial contribution from bias favouring Escitalopram |  | 1.12 (0.93, 1.34) | | 1.13 (0.91, 1.4) | | No evidence of small-study effects | | | Some concerns | |
| **Escitalopram Vs. Sertraline** | 51.11 | 12.28 | 88.88 | Substantial contribution from bias favouring Escitalopram |  | 1.2 (0.96, 1.47) | | 1.14 (0.9, 1.45) | | No evidence of small-study effects | | | Some concerns | |
| **Escitalopram Vs. Venlafaxine** | 43.86 | 26.99 | 87.14 | Substantial contribution from bias favouring Escitalopram |  | 1.13 (0.92, 1.37) | | 1.12 (0.88, 1.42) | | No evidence of small-study effects | | | Some concerns | |
| **Fluoxetine Vs. Fluvoxamine** | 27.12 | 10.24 | 88.08 | Substantial contribution from bias favouring Fluoxetine |  | 1.01 (0.81, 1.26) | | 1.01 (0.76, 1.35) | | No evidence of small-study effects | | | Some concerns | |
| **Fluoxetine Vs. Milnacipran** | 12.29 | 53.29 | 87.75 | Substantial contribution from bias favouring Milnacipran |  | 0.9 (0.7, 1.13) | | 0.91 (0.71, 1.16) | | No evidence of small-study effects | | | Some concerns | |
| **Fluoxetine Vs. Mirtazapine** | 5.92 | 51.83 | 89.50 | Substantial contribution from bias favouring Mirtazapine |  | 0.78 (0.64, 0.94) | | 0.76 (0.62, 0.94) | | No evidence of small-study effects | | | Some concerns | |
| **Fluoxetine Vs. Nefazodone** | 4.59 | 56.53 | 85.34 | Substantial contribution from bias favouring Nefazodone |  | 0.96 (0.66, 1.4) | | 0.94 (0.59, 1.51) | | No evidence of small-study effects | | | Some concerns | |
| **Fluoxetine Vs. Paroxetine** | 10.03 | 0.00 | 50.66 | No substantial contribution from bias |  | 0.84 (0.74, 0.96) | | 0.86 (0.74, 0.99) | | No evidence of small-study effects | | | Low risk | |
| **Fluoxetine Vs. Reboxetine** | 3.18 | 54.99 | 89.57 | Substantial contribution from bias favouring Reboxetine |  | 1.2 (0.88, 1.63) | | 1.21 (0.86, 1.71) | | No evidence of small-study effects | | | Some concerns | |
| **Fluoxetine Vs. Sertraline** | 8.52 | 39.61 | 87.27 | Substantial contribution from bias favouring Sertraline |  | 0.9 (0.76, 1.06) | | 0.87 (0.73, 1.03) | | No evidence of small-study effects | | | Some concerns | |
| **Fluoxetine Vs. Trazodone** | 9.77 | 28.50 | 78.19 | Substantial contribution from bias favouring Trazodone |  | 1.13 (0.86, 1.46) | | 1.1 (0.82, 1.47) | | No evidence of small-study effects | | | Some concerns | |
| **Fluoxetine Vs. Venlafaxine** | 5.80 | 61.26 | 87.08 | Substantial contribution from bias favouring Venlafaxine |  | 0.85 (0.73, 0.97) | | 0.85 (0.71, 1.01) | | No evidence of small-study effects | | | Some concerns | |
| **Fluvoxamine Vs. Milnacipran** | 10.19 | 55.81 | 92.77 | Substantial contribution from bias favouring Milnacipran |  | 0.89 (0.66, 1.18) | | 0.89 (0.63, 1.25) | | No evidence of small-study effects | | | Some concerns | |
| **Fluvoxamine Vs. Mirtazapine** | 6.15 | 56.01 | 93.21 | Substantial contribution from bias favouring Mirtazapine |  | 0.77 (0.6, 1) | | 0.75 (0.55, 1.02) | | No evidence of small-study effects | | | Some concerns | |
| **Fluvoxamine Vs. Paroxetine** | 11.67 | 0.00 | 63.98 | No substantial contribution from bias |  | 0.84 (0.67, 1.04) | | 0.85 (0.63, 1.13) | | No evidence of small-study effects | | | Low risk | |
| **Fluvoxamine Vs. Sertraline** | 10.03 | 34.03 | 90.28 | Substantial contribution from bias favouring Sertraline |  | 0.89 (0.7, 1.14) | | 0.86 (0.63, 1.16) | | No evidence of small-study effects | | | Some concerns | |
| **Fluvoxamine Vs. Venlafaxine** | 9.24 | 40.05 | 90.84 | Substantial contribution from bias favouring Venlafaxine |  | 0.84 (0.66, 1.07) | | 0.84 (0.61, 1.14) | | No evidence of small-study effects | | | Some concerns | |
| **Milnacipran Vs. Paroxetine** | 65.26 | 0.00 | 78.68 | Substantial contribution from bias favouring Milnacipran |  | 0.94 (0.75, 1.18) | | 0.95 (0.74, 1.21) | | No evidence of small-study effects | | | Some concerns | |
| **Milnacipran Vs. Sertraline** | 41.50 | 24.27 | 90.36 | Substantial contribution from bias favouring Milnacipran |  | 1 (0.77, 1.3) | | 0.96 (0.73, 1.26) | | No evidence of small-study effects | | | Some concerns | |
| **Mirtazapine Vs. Paroxetine** | 52.73 | 0.00 | 77.67 | Substantial contribution from bias favouring Mirtazapine |  | 1.08 (0.9, 1.3) | | 1.13 (0.91, 1.39) | | No evidence of small-study effects | | | Some concerns | |
| **Mirtazapine Vs. Sertraline** | 46.60 | 18.64 | 92.18 | Substantial contribution from bias favouring Mirtazapine |  | 1.15 (0.93, 1.43) | | 1.14 (0.89, 1.45) | | No evidence of small-study effects | | | Some concerns | |
| **Mirtazapine Vs. Trazodone** | 45.99 | 13.77 | 85.06 | Substantial contribution from bias favouring Mirtazapine |  | 1.45 (1.08, 1.93) | | 1.44 (1.04, 2) | | No evidence of small-study effects | | | Some concerns | |
| **Mirtazapine Vs. Venlafaxine** | 34.34 | 46.70 | 93.42 | Substantial contribution from bias balanced |  | 1.08 (0.88, 1.32) | | 1.11 (0.88, 1.41) | | No evidence of small-study effects | | | Low risk | |
| **Nefazodone Vs. Paroxetine** | 67.24 | 0.00 | 84.47 | Substantial contribution from bias favouring Nefazodone |  | 0.88 (0.6, 1.28) | | 0.91 (0.57, 1.45) | | No evidence of small-study effects | | | Some concerns | |
| **Nefazodone Vs. Sertraline** | 54.83 | 14.69 | 88.30 | Substantial contribution from bias favouring Nefazodone |  | 0.93 (0.63, 1.38) | | 0.92 (0.57, 1.48) | | No evidence of small-study effects | | | Some concerns | |
| **Paroxetine Vs. Reboxetine** | 0.00 | 47.01 | 80.18 | Substantial contribution from bias favouring Reboxetine |  | 1.43 (1.05, 1.95) | | 1.41 (1, 2) | | No evidence of small-study effects | | | Some concerns | |
| **Paroxetine Vs. Sertraline** | 0.00 | 21.54 | 59.67 | Substantial contribution from bias favouring Sertraline |  | 1.06 (0.91, 1.26) | | 1.01 (0.84, 1.21) | | No evidence of small-study effects | | | Some concerns | |
| **Paroxetine Vs. Trazodone** | 0.00 | 19.20 | 46.27 | Substantial contribution from bias favouring Trazodone |  | 1.34 (1.03, 1.73) | | 1.28 (0.95, 1.72) | | No evidence of small-study effects | | | Some concerns | |
| **Paroxetine Vs. Venlafaxine** | 0.00 | 41.77 | 68.14 | Substantial contribution from bias favouring Venlafaxine |  | 1 (0.86, 1.17) | | 0.99 (0.82, 1.19) | | No evidence of small-study effects | | | Some concerns | |
| **Reboxetine Vs. Venlafaxine** | 32.69 | 46.58 | 92.60 | Substantial contribution from bias balanced |  | 0.7 (0.52, 0.96) | | 0.7 (0.48, 1.01) | | No evidence of small-study effects | | | Low risk | |
| **Sertraline Vs. Trazodone** | 32.22 | 18.27 | 82.27 | Substantial contribution from bias balanced |  | 1.26 (0.95, 1.65) | | 1.27 (0.92, 1.74) | | No evidence of small-study effects | | | Low risk | |
| **Sertraline Vs. Venlafaxine** | 19.77 | 47.68 | 90.58 | Substantial contribution from bias favouring Venlafaxine |  | 0.94 (0.79, 1.13) | | 0.98 (0.79, 1.21) | | No evidence of small-study effects | | | Some concerns | |
| **Trazodone Vs. Venlafaxine** | 18.21 | 44.02 | 84.15 | Substantial contribution from bias favouring Venlafaxine |  | 0.75 (0.57, 0.98) | | 0.77 (0.56, 1.06) | | No evidence of small-study effects | | | Some concerns | |
| **Venlafaxine Vs. Vortioxetine** | 0.00 | 0.00 | 0.00 | No substantial contribution from bias |  | 0.72 (0.43, 1.19) | | 0.71 (0.43, 1.19) | | No evidence of small-study effects | | | Low risk | |
| **Only indirect** | | | | | | | | | | | | | | |
| **Amitriptyline Vs. Bupropion** | 0.00 | 0.00 | 59.59 | No substantial contribution from bias | Undetected bias | 0.91 (0.62, 1.33) | | 0.93 (0.61, 1.39) | | | No evidence of small-study effects | | | Low risk |
| **Amitriptyline Vs. Nefazodone** | 0.00 | 45.16 | 80.12 | Substantial contribution from bias favouring Nefazodone | Suspected bias favouring Nefazodone | 1.2 (0.81, 1.78) | | 1.22 (0.74, 1.99) | | | No evidence of small-study effects | | | High risk |
| **Agomelatine Vs. Amitriptyline** | 0.00 | 0.00 | 43.12 | No substantial contribution from bias | Undetected bias | 0.96 (0.75, 1.22) | | 0.91 (0.68, 1.2) | | | No evidence of small-study effects | | | Low risk |
| **Agomelatine Vs. Bupropion** | 0.00 | 0.00 | 31.49 | No substantial contribution from bias | Undetected bias | 0.87 (0.58, 1.3) | | 0.85 (0.55, 1.3) | | | No evidence of small-study effects | | | Low risk |
| **Agomelatine Vs. Citalopram** | 0.00 | 13.86 | 57.14 | No substantial contribution from bias | Undetected bias | 1.13 (0.87, 1.47) | | 1.15 (0.86, 1.54) | | | No evidence of small-study effects | | | Low risk |
| **Agomelatine Vs. Clomipramine** | 0.00 | 0.00 | 42.80 | No substantial contribution from bias | Undetected bias | 1.2 (0.92, 1.57) | | 1.12 (0.8, 1.57) | | | No evidence of small-study effects | | | Low risk |
| **Agomelatine Vs. Fluvoxamine** | 0.00 | 7.11 | 56.17 | No substantial contribution from bias | Undetected bias | 1.2 (0.91, 1.61) | | 1.19 (0.83, 1.69) | | | No evidence of small-study effects | | | Low risk |
| **Agomelatine Vs. Milnacipran** | 0.00 | 37.32 | 59.77 | Substantial contribution from bias favouring Milnacipran | Undetected bias | 1.07 (0.8, 1.45) | | 1.06 (0.77, 1.47) | | | No evidence of small-study effects | | | Some concerns |
| **Agomelatine Vs. Mirtazapine** | 0.00 | 34.39 | 57.36 | Substantial contribution from bias favouring Mirtazapine | Undetected bias | 0.93 (0.72, 1.21) | | 0.89 (0.67, 1.19) | | | No evidence of small-study effects | | | Some concerns |
| **Agomelatine Vs. Nefazodone** | 0.00 | 39.26 | 58.71 | Substantial contribution from bias favouring Nefazodone | Undetected bias | 1.15 (0.76, 1.74) | | 1.11 (0.67, 1.84) | | | No evidence of small-study effects | | | Some concerns |
| **Agomelatine Vs. Reboxetine** | 0.00 | 34.34 | 56.21 | Substantial contribution from bias favouring Reboxetine | Undetected bias | 1.44 (1.02, 2.05) | | 1.42 (0.96, 2.13) | | | No evidence of small-study effects | | | Some concerns |
| **Agomelatine Vs. Sertraline** | 0.00 | 17.88 | 49.41 | Substantial contribution from bias favouring Sertraline | Undetected bias | 1.07 (0.85, 1.37) | | 1.02 (0.78, 1.33) | | | No evidence of small-study effects | | | Some concerns |
| **Agomelatine Vs. Trazodone** | 0.00 | 15.08 | 45.35 | Substantial contribution from bias favouring Trazodone | Undetected bias | 1.35 (0.98, 1.86) | | 1.29 (0.9, 1.84) | | | No evidence of small-study effects | | | Some concerns |
| **Agomelatine Vs. Vortioxetine** | 0.00 | 0.00 | 25.49 | No substantial contribution from bias | Undetected bias | 0.72 (0.42, 1.25) | | 0.71 (0.4, 1.25) | | | No evidence of small-study effects | | | Low risk |
| **Amitriptyline Vs. Citalopram** | 0.00 | 19.30 | 85.93 | Substantial contribution from bias favouring Citalopram | Suspected bias favouring Citalopram | 1.18 (0.93, 1.48) | | 1.27 (0.97, 1.67) | | | No evidence of small-study effects | | | High risk |
| **Amitriptyline Vs. Clomipramine** | 0.00 | 0.00 | 65.17 | No substantial contribution from bias | Undetected bias | 1.25 (0.99, 1.59) | | 1.24 (0.91, 1.67) | | | No evidence of small-study effects | | | Low risk |
| **Amitriptyline Vs. Duloxetine** | 0.00 | 33.46 | 71.39 | Substantial contribution from bias favouring Duloxetine | Suspected bias favouring Duloxetine | 1.1 (0.85, 1.42) | | 1.13 (0.82, 1.52) | | | No evidence of small-study effects | | | High risk |
| **Amitriptyline Vs. Escitalopram** | 0.00 | 34.23 | 79.51 | Substantial contribution from bias favouring Escitalopram | Suspected bias favouring Escitalopram | 0.94 (0.74, 1.18) | | 0.98 (0.75, 1.27) | | | No evidence of small-study effects | | | High risk |
| **Amitriptyline Vs. Mirtazapine** | 0.00 | 39.83 | 84.06 | Substantial contribution from bias favouring Mirtazapine | Suspected bias favouring Mirtazapine | 0.98 (0.78, 1.21) | | 0.98 (0.76, 1.27) | | | No evidence of small-study effects | | | High risk |
| **Amitriptyline Vs. Reboxetine** | 0.00 | 35.24 | 83.10 | Substantial contribution from bias favouring Reboxetine | Suspected bias favouring Reboxetine | 1.5 (1.07, 2.09) | | 1.56 (1.07, 2.29) | | | No evidence of small-study effects | | | High risk |
| **Amitriptyline Vs. Vortioxetine** | 0.00 | 0.00 | 54.86 | No substantial contribution from bias | Undetected bias | 0.76 (0.44, 1.28) | | 0.78 (0.44, 1.35) | | | No evidence of small-study effects | | | Low risk |
| **Bupropion Vs. Citalopram** | 0.00 | 15.87 | 76.28 | Substantial contribution from bias favouring Citalopram | Suspected bias favouring Citalopram | 1.29 (0.88, 1.93) | | 1.36 (0.89, 2.09) | | | No evidence of small-study effects | | | High risk |
| **Bupropion Vs. Clomipramine** | 0.00 | 0.00 | 59.39 | No substantial contribution from bias | Undetected bias | 1.38 (0.92, 2.07) | | 1.33 (0.85, 2.09) | | | No evidence of small-study effects | | | Low risk |
| **Bupropion Vs. Duloxetine** | 0.00 | 33.99 | 67.24 | Substantial contribution from bias favouring Duloxetine | Suspected bias favouring Duloxetine | 1.21 (0.81, 1.82) | | 1.21 (0.78, 1.9) | | | No evidence of small-study effects | | | High risk |
| **Bupropion Vs. Escitalopram** | 0.00 | 32.49 | 71.24 | Substantial contribution from bias favouring Escitalopram | Suspected bias favouring Escitalopram | 1.03 (0.7, 1.53) | | 1.05 (0.7, 1.61) | | | No evidence of small-study effects | | | High risk |
| **Bupropion Vs. Fluvoxamine** | 0.00 | 7.41 | 71.13 | No substantial contribution from bias | Suspected bias favouring Fluvoxamine | 1.38 (0.92, 2.08) | | 1.41 (0.89, 2.25) | | | No evidence of small-study effects | | | Low risk |
| **Bupropion Vs. Milnacipran** | 0.00 | 37.49 | 71.60 | Substantial contribution from bias favouring Milnacipran | Suspected bias favouring Milnacipran | 1.23 (0.81, 1.88) | | 1.26 (0.82, 1.97) | | | No evidence of small-study effects | | | High risk |
| **Bupropion Vs. Mirtazapine** | 0.00 | 35.52 | 75.02 | Substantial contribution from bias favouring Mirtazapine | Suspected bias favouring Mirtazapine | 1.07 (0.72, 1.59) | | 1.06 (0.7, 1.61) | | | No evidence of small-study effects | | | High risk |
| **Bupropion Vs. Nefazodone** | 0.00 | 38.92 | 71.05 | Substantial contribution from bias favouring Nefazodone | Suspected bias favouring Nefazodone | 1.32 (0.8, 2.23) | | 1.31 (0.73, 2.39) | | | No evidence of small-study effects | | | High risk |
| **Bupropion Vs. Reboxetine** | 0.00 | 33.10 | 74.28 | Substantial contribution from bias favouring Reboxetine | Suspected bias favouring Reboxetine | 1.65 (1.03, 2.64) | | 1.68 (1.02, 2.79) | | | No evidence of small-study effects | | | High risk |
| **Bupropion Vs. Vortioxetine** | 0.00 | 0.00 | 45.16 | No substantial contribution from bias | Undetected bias | 0.83 (0.45, 1.53) | | 0.84 (0.45, 1.61) | | | No evidence of small-study effects | | | Low risk |
| **Citalopram Vs. Duloxetine** | 10.33 | 32.46 | 84.33 | Substantial contribution from bias favouring Duloxetine | Suspected bias favouring Duloxetine | 0.93 (0.71, 1.23) | | 0.89 (0.64, 1.21) | | | No evidence of small-study effects | | | High risk |
| **Citalopram Vs. Milnacipran** | 17.42 | 34.04 | 94.01 | Substantial contribution from bias favouring Milnacipran | Suspected bias favouring Milnacipran | 0.95 (0.72, 1.26) | | 0.92 (0.67, 1.25) | | | No evidence of small-study effects | | | High risk |
| **Citalopram Vs. Nefazodone** | 19.65 | 36.88 | 93.23 | Substantial contribution from bias favouring Nefazodone | Suspected bias favouring Nefazodone | 1.02 (0.67, 1.55) | | 0.96 (0.58, 1.6) | | | No evidence of small-study effects | | | High risk |
| **Citalopram Vs. Paroxetine** | 17.53 | 0.00 | 75.07 | Substantial contribution from bias favouring Citalopram | Undetected bias | 0.89 (0.73, 1.1) | | 0.88 (0.69, 1.1) | | | No evidence of small-study effects | | | Some concerns |
| **Citalopram Vs. Trazodone** | 18.15 | 14.79 | 86.64 | Substantial contribution from bias balanced | Suspected bias favouring Trazodone | 1.19 (0.88, 1.63) | | 1.12 (0.78, 1.58) | | | No evidence of small-study effects | | | Low risk |
| **Citalopram Vs. Vortioxetine** | 11.59 | 0.00 | 64.05 | No substantial contribution from bias | Undetected bias | 0.64 (0.37, 1.12) | | 0.62 (0.35, 1.07) | | | No evidence of small-study effects | | | Low risk |
| **Clomipramine Vs. Duloxetine** | 0.00 | 34.20 | 71.91 | Substantial contribution from bias favouring Duloxetine | Suspected bias favouring Duloxetine | 0.88 (0.66, 1.16) | | 0.91 (0.63, 1.29) | | | No evidence of small-study effects | | | High risk |
| **Clomipramine Vs. Escitalopram** | 0.00 | 35.02 | 79.12 | Substantial contribution from bias favouring Escitalopram | Suspected bias favouring Escitalopram | 0.75 (0.58, 0.96) | | 0.79 (0.58, 1.09) | | | No evidence of small-study effects | | | High risk |
| **Clomipramine Vs. Mirtazapine** | 0.00 | 40.40 | 83.31 | Substantial contribution from bias favouring Mirtazapine | Suspected bias favouring Mirtazapine | 0.78 (0.6, 1.01) | | 0.79 (0.58, 1.1) | | | No evidence of small-study effects | | | High risk |
| **Clomipramine Vs. Nefazodone** | 0.00 | 43.59 | 80.17 | Substantial contribution from bias favouring Nefazodone | Suspected bias favouring Nefazodone | 0.96 (0.63, 1.47) | | 0.99 (0.59, 1.65) | | | No evidence of small-study effects | | | High risk |
| **Clomipramine Vs. Reboxetine** | 0.00 | 34.10 | 82.26 | Substantial contribution from bias favouring Reboxetine | Suspected bias favouring Reboxetine | 1.2 (0.84, 1.72) | | 1.27 (0.83, 1.94) | | | No evidence of small-study effects | | | High risk |
| **Clomipramine Vs. Vortioxetine** | 0.00 | 0.00 | 53.72 | No substantial contribution from bias | Undetected bias | 0.6 (0.34, 1.04) | | 0.63 (0.35, 1.13) | | | No evidence of small-study effects | | | Low risk |
| **Duloxetine Vs. Fluoxetine** | 34.34 | 4.70 | 72.80 | Substantial contribution from bias favouring Duloxetine | Suspected bias favouring Duloxetine | 1.14 (0.91, 1.44) | | 1.14 (0.88, 1.5) | | | No evidence of small-study effects | | | High risk |
| **Duloxetine Vs. Fluvoxamine** | 28.26 | 7.24 | 80.04 | Substantial contribution from bias favouring Duloxetine | Suspected bias favouring Duloxetine | 1.14 (0.85, 1.55) | | 1.16 (0.81, 1.7) | | | No evidence of small-study effects | | | High risk |
| **Duloxetine Vs. Milnacipran** | 30.60 | 35.51 | 87.40 | Substantial contribution from bias balanced | Suspected bias favouring Duloxetine | 1.02 (0.75, 1.4) | | 1.03 (0.74, 1.48) | | | No evidence of small-study effects | | | Low risk |
| **Duloxetine Vs. Mirtazapine** | 33.66 | 30.97 | 87.52 | Substantial contribution from bias balanced | Suspected bias favouring Duloxetine | 0.89 (0.67, 1.17) | | 0.87 (0.64, 1.2) | | | No evidence of small-study effects | | | Low risk |
| **Duloxetine Vs. Nefazodone** | 32.14 | 37.16 | 89.33 | Substantial contribution from bias balanced | Suspected bias favouring Duloxetine | 1.09 (0.71, 1.68) | | 1.08 (0.64, 1.83) | | | No evidence of small-study effects | | | Low risk |
| **Duloxetine Vs. Reboxetine** | 34.10 | 29.65 | 87.96 | Substantial contribution from bias balanced | Suspected bias favouring Duloxetine | 1.37 (0.94, 1.97) | | 1.39 (0.92, 2.1) | | | No evidence of small-study effects | | | Low risk |
| **Duloxetine Vs. Sertraline** | 33.88 | 13.45 | 78.83 | Substantial contribution from bias favouring Duloxetine | Suspected bias favouring Duloxetine | 1.02 (0.79, 1.31) | | 0.99 (0.75, 1.35) | | | No evidence of small-study effects | | | High risk |
| **Duloxetine Vs. Trazodone** | 32.28 | 12.06 | 72.96 | Substantial contribution from bias favouring Duloxetine | Suspected bias favouring Duloxetine | 1.28 (0.92, 1.78) | | 1.26 (0.86, 1.85) | | | No evidence of small-study effects | | | High risk |
| **Duloxetine Vs. Vortioxetine** | 28.91 | 0.00 | 50.96 | Substantial contribution from bias favouring Duloxetine | Undetected bias | 0.69 (0.39, 1.2) | | 0.69 (0.39, 1.24) | | | No evidence of small-study effects | | | Some concerns |
| **Escitalopram Vs. Fluvoxamine** | 33.32 | 6.52 | 88.02 | Substantial contribution from bias favouring Escitalopram | Suspected bias favouring Escitalopram | 1.34 (1.02, 1.75) | | 1.34 (0.96, 1.87) | | | No evidence of small-study effects | | | High risk |
| **Escitalopram Vs. Milnacipran** | 29.80 | 35.11 | 90.43 | Substantial contribution from bias balanced | Suspected bias favouring Escitalopram | 1.2 (0.91, 1.58) | | 1.2 (0.88, 1.63) | | | No evidence of small-study effects | | | Low risk |
| **Escitalopram Vs. Mirtazapine** | 37.14 | 35.93 | 93.24 | Substantial contribution from bias balanced | Suspected bias favouring Escitalopram | 1.04 (0.81, 1.33) | | 1.01 (0.77, 1.31) | | | No evidence of small-study effects | | | Low risk |
| **Escitalopram Vs. Nefazodone** | 33.61 | 38.75 | 91.10 | Substantial contribution from bias balanced | Suspected bias favouring Escitalopram | 1.28 (0.84, 1.92) | | 1.24 (0.76, 2.06) | | | No evidence of small-study effects | | | Low risk |
| **Escitalopram Vs. Reboxetine** | 37.07 | 34.58 | 92.58 | Substantial contribution from bias balanced | Suspected bias favouring Escitalopram | 1.61 (1.15, 2.22) | | 1.6 (1.09, 2.34) | | | No evidence of small-study effects | | | Low risk |
| **Escitalopram Vs. Trazodone** | 33.55 | 13.64 | 80.15 | Substantial contribution from bias favouring Escitalopram | Suspected bias favouring Escitalopram | 1.51 (1.1, 2.03) | | 1.45 (1.03, 2.04) | | | No evidence of small-study effects | | | High risk |
| **Escitalopram Vs. Vortioxetine** | 27.19 | 0.00 | 58.05 | Substantial contribution from bias favouring Escitalopram | Undetected bias | 0.81 (0.46, 1.39) | | 0.8 (0.45, 1.38) | | | No evidence of small-study effects | | | Some concerns |
| **Fluoxetine Vs. Vortioxetine** | 3.97 | 0.00 | 51.99 | No substantial contribution from bias | Undetected bias | 0.6 (0.35, 1.01) | | 0.61 (0.35, 1.03) | | | No evidence of small-study effects | | | Low risk |
| **Fluvoxamine Vs. Nefazodone** | 7.62 | 38.15 | 89.41 | Substantial contribution from bias favouring Nefazodone | Suspected bias favouring Nefazodone | 0.95 (0.62, 1.46) | | 0.93 (0.55, 1.58) | | | No evidence of small-study effects | | | High risk |
| **Fluvoxamine Vs. Reboxetine** | 7.57 | 31.53 | 91.65 | Substantial contribution from bias favouring Reboxetine | Suspected bias favouring Reboxetine | 1.2 (0.83, 1.7) | | 1.2 (0.77, 1.83) | | | No evidence of small-study effects | | | High risk |
| **Fluvoxamine Vs. Trazodone** | 11.64 | 19.55 | 83.15 | Substantial contribution from bias balanced | Suspected bias favouring Fluvoxamine | 1.12 (0.81, 1.54) | | 1.09 (0.73, 1.6) | | | No evidence of small-study effects | | | Low risk |
| **Fluvoxamine Vs. Vortioxetine** | 6.50 | 0.00 | 63.24 | No substantial contribution from bias | Undetected bias | 0.6 (0.34, 1.05) | | 0.6 (0.33, 1.07) | | | No evidence of small-study effects | | | Low risk |
| **Milnacipran Vs. Mirtazapine** | 41.56 | 38.12 | 95.92 | Substantial contribution from bias balanced | Suspected bias favouring Milnacipran | 0.87 (0.66, 1.15) | | 0.84 (0.62, 1.13) | | | No evidence of small-study effects | | | Low risk |
| **Milnacipran Vs. Nefazodone** | 41.49 | 41.49 | 95.90 | Substantial contribution from bias balanced | Suspected bias favouring Milnacipran | 1.07 (0.7, 1.63) | | 1.04 (0.62, 1.75) | | | No evidence of small-study effects | | | Low risk |
| **Milnacipran Vs. Reboxetine** | 37.88 | 32.84 | 95.93 | Substantial contribution from bias balanced | Suspected bias favouring Reboxetine | 1.34 (0.93, 1.93) | | 1.34 (0.89, 2.02) | | | No evidence of small-study effects | | | Low risk |
| **Milnacipran Vs. Trazodone** | 43.26 | 20.28 | 83.86 | Substantial contribution from bias favouring Milnacipran | Suspected bias favouring Trazodone | 1.26 (0.9, 1.74) | | 1.21 (0.83, 1.75) | | | No evidence of small-study effects | | | Some concerns |
| **Milnacipran Vs. Venlafaxine** | 39.54 | 33.78 | 91.16 | Substantial contribution from bias balanced | Suspected bias favouring Venlafaxine | 0.94 (0.73, 1.21) | | 0.94 (0.7, 1.25) | | | No evidence of small-study effects | | | Low risk |
| **Milnacipran Vs. Vortioxetine** | 27.96 | 0.00 | 65.29 | Substantial contribution from bias favouring Milnacipran | Undetected bias | 0.67 (0.38, 1.17) | | 0.67 (0.37, 1.19) | | | No evidence of small-study effects | | | Some concerns |
| **Mirtazapine Vs. Nefazodone** | 37.50 | 41.22 | 95.80 | Substantial contribution from bias balanced | Suspected bias favouring Nefazodone | 1.23 (0.81, 1.85) | | 1.24 (0.76, 2.04) | | | No evidence of small-study effects | | | Low risk |
| **Mirtazapine Vs. Reboxetine** | 37.70 | 36.38 | 98.28 | Substantial contribution from bias balanced | Suspected bias favouring Reboxetine | 1.54 (1.09, 2.17) | | 1.59 (1.08, 2.35) | | | No evidence of small-study effects | | | Low risk |
| **Mirtazapine Vs. Vortioxetine** | 23.93 | 0.00 | 62.17 | Substantial contribution from bias favouring Mirtazapine | Undetected bias | 0.77 (0.45, 1.33) | | 0.8 (0.45, 1.39) | | | No evidence of small-study effects | | | Some concerns |
| **Nefazodone Vs. Reboxetine** | 39.74 | 34.99 | 96.13 | Substantial contribution from bias balanced | Suspected bias favouring Reboxetine | 1.25 (0.79, 2) | | 1.29 (0.72, 2.28) | | | No evidence of small-study effects | | | Low risk |
| **Nefazodone Vs. Trazodone** | 41.23 | 15.79 | 81.05 | Substantial contribution from bias favouring Nefazodone | Suspected bias favouring Nefazodone | 1.17 (0.75, 1.85) | | 1.17 (0.68, 1.98) | | | No evidence of small-study effects | | | High risk |
| **Nefazodone Vs. Venlafaxine** | 40.54 | 34.41 | 89.33 | Substantial contribution from bias balanced | Suspected bias favouring Venlafaxine | 0.88 (0.59, 1.31) | | 0.9 (0.56, 1.46) | | | No evidence of small-study effects | | | Low risk |
| **Nefazodone Vs. Vortioxetine** | 28.50 | 0.00 | 63.36 | Substantial contribution from bias favouring Nefazodone | Undetected bias | 0.63 (0.33, 1.2) | | 0.64 (0.32, 1.29) | | | No evidence of small-study effects | | | Some concerns |
| **Paroxetine Vs. Vortioxetine** | 0.00 | 0.00 | 44.91 | No substantial contribution from bias | Undetected bias | 0.72 (0.42, 1.21) | | 0.71 (0.41, 1.2) | | | No evidence of small-study effects | | | Low risk |
| **Reboxetine Vs. Sertraline** | 35.19 | 18.50 | 91.15 | Substantial contribution from bias favouring Reboxetine | Suspected bias favouring Reboxetine | 0.75 (0.54, 1.04) | | 0.72 (0.49, 1.04) | | | No evidence of small-study effects | | | High risk |
| **Reboxetine Vs. Trazodone** | 32.36 | 15.20 | 83.36 | Substantial contribution from bias favouring Reboxetine | Suspected bias favouring Reboxetine | 0.94 (0.63, 1.39) | | 0.91 (0.58, 1.41) | | | No evidence of small-study effects | | | High risk |
| **Reboxetine Vs. Vortioxetine** | 22.99 | 0.00 | 61.69 | Substantial contribution from bias favouring Reboxetine | Undetected bias | 0.5 (0.27, 0.92) | | 0.5 (0.27, 0.93) | | | No evidence of small-study effects | | | Some concerns |
| **Sertraline Vs. Vortioxetine** | 13.42 | 0.00 | 59.01 | No substantial contribution from bias | Undetected bias | 0.67 (0.39, 1.15) | | 0.7 (0.4, 1.2) | | | No evidence of small-study effects | | | Low risk |
| **Trazodone Vs. Vortioxetine** | 12.50 | 0.00 | 56.54 | No substantial contribution from bias | Undetected bias | 0.53 (0.3, 0.95) | | 0.55 (0.3, 1) | | | No evidence of small-study effects | | | Low risk |
